# Rapid and portable reverse-transcription quantitative PCR assays for Bundibugyo ebolavirus detection

**DOI:** 10.64898/2026.08.17.26360605

**Authors:** Kyle McMahon, Stella Nielsen, Hannah Knoll, Resham Talwar, Davina Thompson, Colby Wilkason, Al Ozonoff, Elyse Stachler, Pardis C. Sabeti

## Abstract

The 2026 Bundibugyo ebolavirus (BDBV) outbreak underscores the need for rapidly deployable molecular diagnostics. We developed and analytically validated reverse-transcription quantitative PCR assays detecting BDBV, Zaire ebolavirus, and Sudan ebolavirus. The platform includes a BDBV singleplex assay, a duplex assay with a human internal control, a four-target multiplex assay for ebolavirus differentiation, and a probe-free SYBR Green assay. We adapted the assays to a portable qPCR instrument, reducing runtime from 65 to 35 minutes, and validated lyophilized reagents to reduce cold-chain requirements. All TaqMan formats achieved a 95% limit of detection of 5 copies per reaction across instruments and reagent types; the SYBR Green assay achieved 50 copies per reaction. The assays detected viral RNA in contrived clinical samples without cross-reactivity among ebolavirus species tested. We shared the protocols in real time through Ampliphi (https://www.ampliphi.bio), a new open-access platform for rapidly disseminating diagnostic assays, and through protocol.io.

## Introduction

On May 5, 2026, the Ministry of Health of the Democratic Republic of the Congo (DRC) alerted the World Health Organization (WHO) to an outbreak of unknown etiology in the Mongbwalu Health Zone, Ituri Province, following the deaths of four healthcare workers (*1*). On May 15, 2026, rapid response teams confirmed the outbreak as Ebolavirus Disease (EVD) caused by the Bundibugyo virus (BDBV) (*2*). Two days later, the WHO declared the outbreak a Public Health Emergency of International Concern (PHEIC) due to the geographic spread within the DRC and into Uganda, and the rapidly rising caseload (*3*). As of August 8, 2026, the two countries had reported 4,229 laboratory-confirmed cases and 1,918 confirmed deaths (*4*). The speed and scale of the outbreak underscored the need for reliable diagnostic tools that can be rapidly deployed.

BDBV is one of four ebolavirus species known to cause EVD in humans, but it has caused far fewer recognized outbreaks than Zaire (Z-EBOV) and Sudan (S-EBOV) ebolaviruses. These viruses belong to the genus *Orthoebolavirus* within the family *Filoviridae*, which is composed of six genetically distinct species of filamentous, linear, non-segmented, negative sense, single-stranded RNA viruses (*5*,*6*). Taï Forest (CI-EBOV) ebolavirus is the fourth species known to cause disease in humans, while the two remaining species Reston (RESTV) and Bombali (BOMV) ebolaviruses circulate only in animal hosts (*5*,*7*). The 2026 outbreak is only the third recognized outbreak attributed to BDBV and the 17th Ebola outbreak reported in the DRC since 1976. Most EVD outbreaks, cases, and deaths reported from 1976 through 2025 were caused by Z-EBOV, followed by S-EBOV, BDBV, and CI-EBOV (*8*).

Since Z-EBOV has caused the largest number of EVD outbreaks and highest overall disease burden, diagnostic development has focused predominantly on Z-EBOV, with fewer validated options for S-EBOV and BDBV (*9*,*10*). The 2014 West Africa Ebola epidemic accelerated the development and commercialization of molecular tests and vaccines targeting Z-EBOV (*11*,*12*). However, genetic divergence among ebolavirus species means that assays designed for Z-EBOV and S-EBOV may not reliably detect BDBV (*13*). Commercial reverse-transcription quantitative PCR (RT-qPCR) assays for BDBV are available, but their proprietary sequences cannot be readily evaluated against newly generated outbreak genomes (*9*). Heavy demand during an outbreak may also strain commercial supply chains. These limitations create a need for open, sequence-transparent assays that laboratories can evaluate, reproduce, and adapt as an outbreak evolves.

The limited availability of open, non-proprietary BDBV assays motivated us to develop RT-qPCR assays designed against historical BDBV genomes and confirmed against sequences from the circulating outbreak strain. We validated four complementary assay formats suited for different operational needs: (1) a sensitive probe-based BDBV singleplex assay, (2) a duplex assay combining BDBV detection with a human internal control, (3) a four-target multiplex assay that differentiates BDBV, Z-EBOV, and S-EBOV, combined with a human internal control, and (4) a probe-free SYBR Green BDBV assay for settings where probe synthesis or supply is constrained.

To support decentralized testing, we adapted the BDBV singleplex and duplex TaqMan assays to the Mic qPCR cycler, a compact four-channel rotor-based instrument with rapid cycling capabilities that can operate using an external battery (*14*). We also evaluated lyophilized RT-qPCR reagents to reduce cold chain requirements. Together, the portable qPCR instrument, shortened cycling protocol, and shelf-stable reagents provide a flexible workflow for rapid BDBV testing in outbreak-response settings. We shared the resulting assay designs and protocols in real time through Ampliphi, a new open-access platform for rapidly disseminating diagnostic assays.

## Results

### *In silico* sensitivity and specificity

We designed RT-qPCR primers and probes to detect BDBV, Z-EBOV, and S-EBOV and adapted a previously published human mitochondrial circular DNA assay (mcirDNA) as an internal control (Table 1) (*15*). The BDBV assay targets a 103 bp segment of the large polymerase (L) gene, a highly conserved region which encodes the viral RNA-dependent RNA polymerase. At the time of design, the BDBV primers and probes demonstrated 100% sequence identity to all 26 complete historical BDBV genomes available from NCBI Virus (Figure A1). After sequences from this outbreak became available, the assay showed 100% sequence identity across all publicly available genomes (n=533 with complete sequencing data in the amplicon region as of August 10th, 2026) (Figure A2). The BDBV, Z-EBOV, and S-EBOV assays each showed complete predicted coverage of the intended targets, with no predicted cross-reactivity to off-target species.

**Table 1:**
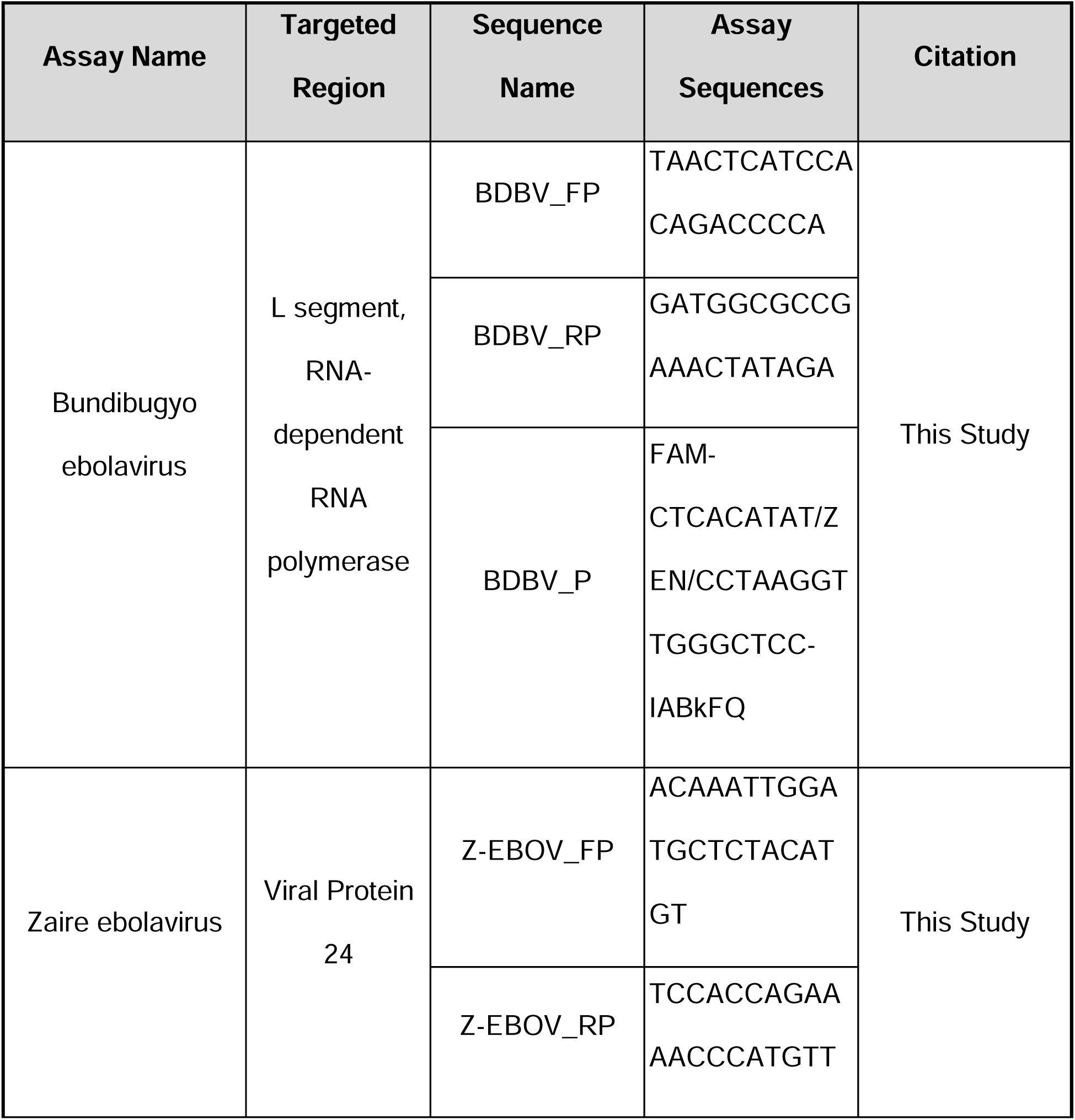
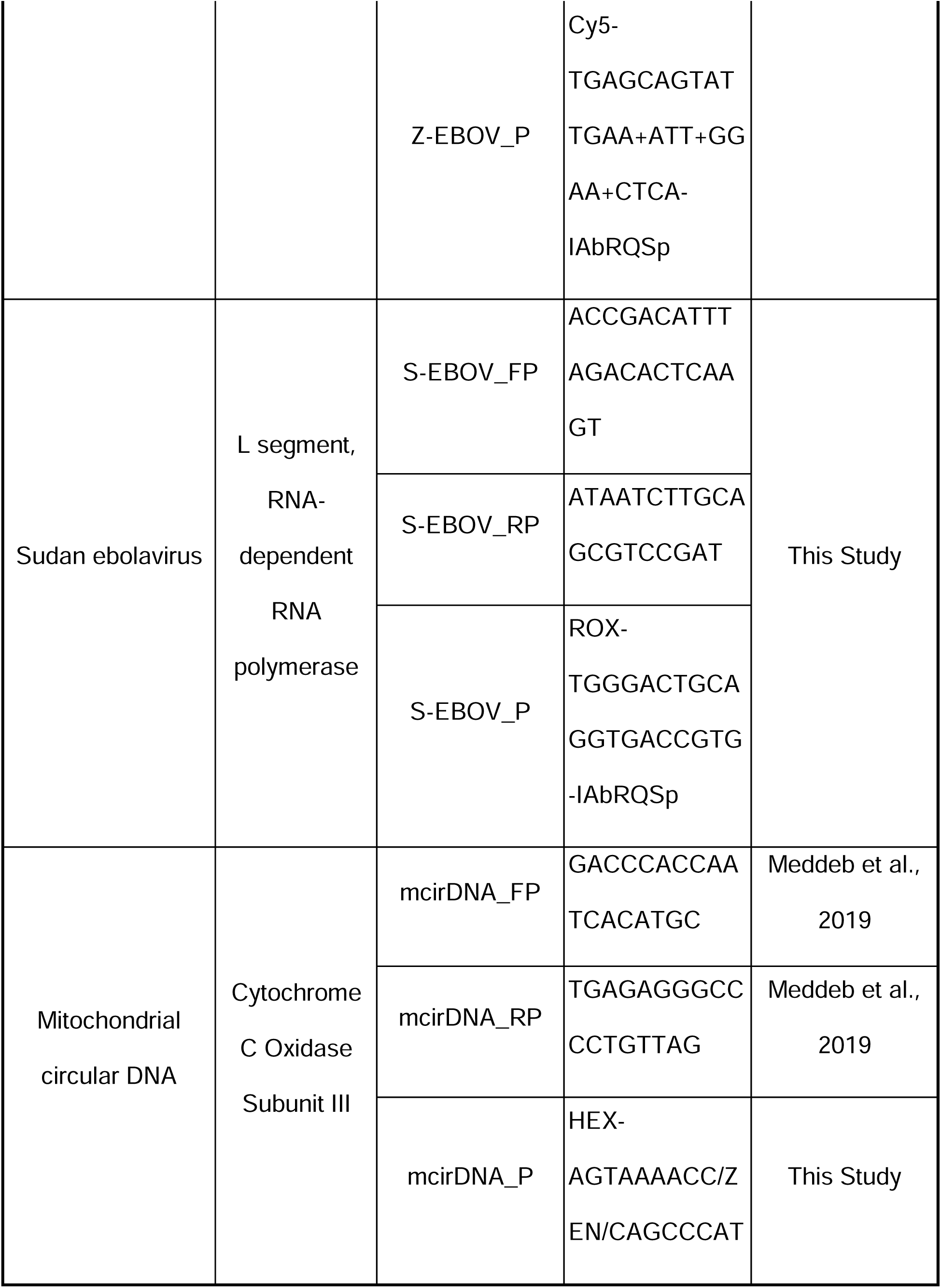
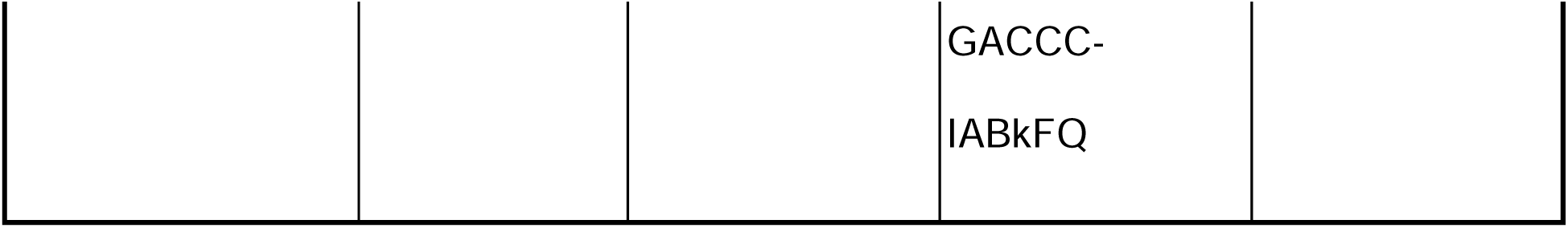
Primer and probe sequences for Bundibugyo ebolavirus (BDBV), Zaire ebolavirus (Z-EBOV), Sudan ebolavirus (S-EBOV) and mitochondrial circular DNA (mcirDNA). The table lists the target region, oligonucleotide name, sequence, and source for each primer and probe used in this study. Locked nucleic acid bases in the Z-EBOV probe are indicated by a plus sign preceding the modified nucleotide.

### Analytical performance under standard RT-qPCR conditions

We evaluated three TaqMan assay formats: (1) a BDBV singleplex assay, (2) a duplex assay detecting BDBV (FAM) and the mcirDNA internal control (HEX), and (3) a four-target multiplex assay detecting BDBV (FAM), Z-EBOV (Cy5), S-EBOV (ROX), and mcirDNA (HEX). We assessed assay performance using synthetic RNA or DNA gene fragments at known concentrations and contrived clinical samples, defined here as whole viral RNA spiked into normalized human plasma background. We first optimized each assay in singleplex format and selected final concentrations of 400nM forward and reverse primer, and 200nM probe based on amplification efficiency between 90–110%, linearity, and observed sensitivity (Figure A3).

The BDBV and mcirDNA assays achieved amplification efficiencies of 98.3– 102.3% in singleplex and duplex formats, with R^2^ values ≥ 0.99 (Figure 1). In four-target multiplex format, all assays achieved efficiencies of 90.8–99.0% and R^2^ values ≥ 0.99 (Figure 2). All TaqMan assay formats achieved an LOD_95_ of 5 copies per reaction, indicating no loss of analytical sensitivity with increasing multiplexing complexity (Table 2, full version available as Table A1).

**Figure 1:**
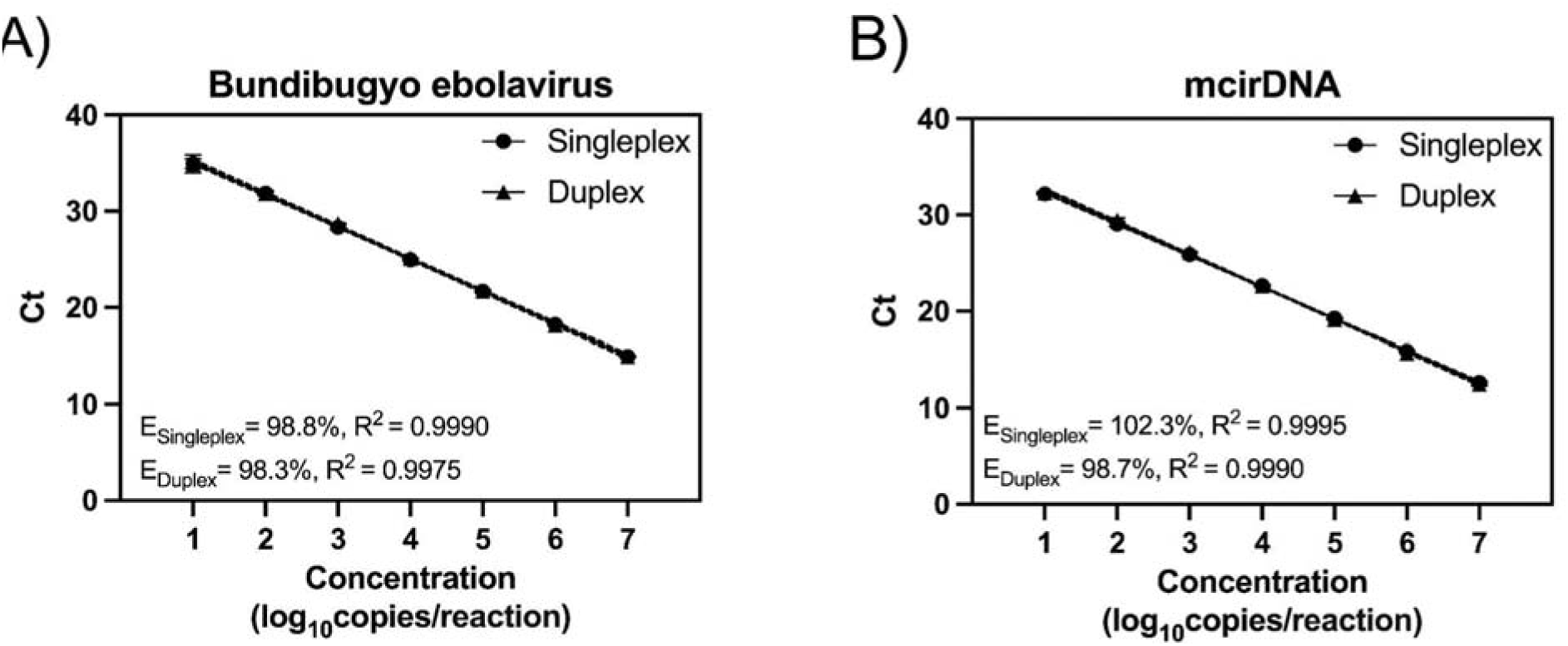
Standard curves for Bundibugyo ebolavirus (BDBV) and mitochondrial circular DNA (mcirDNA) TaqMan RT-qPCR assays in singleplex and duplex formats. We generated standard curves for A) BDBV and B) the mcirDNA internal control assays using serial dilutions of synthetic RNA or DNA gene fragments. Data points show the mean and standard deviation of triplicate reactions. Solid lines indicate simple linear regressions, and dotted lines indicate the corresponding 95% confidence intervals. E denotes the RT-qPCR amplification efficiency.

**Figure 2:**
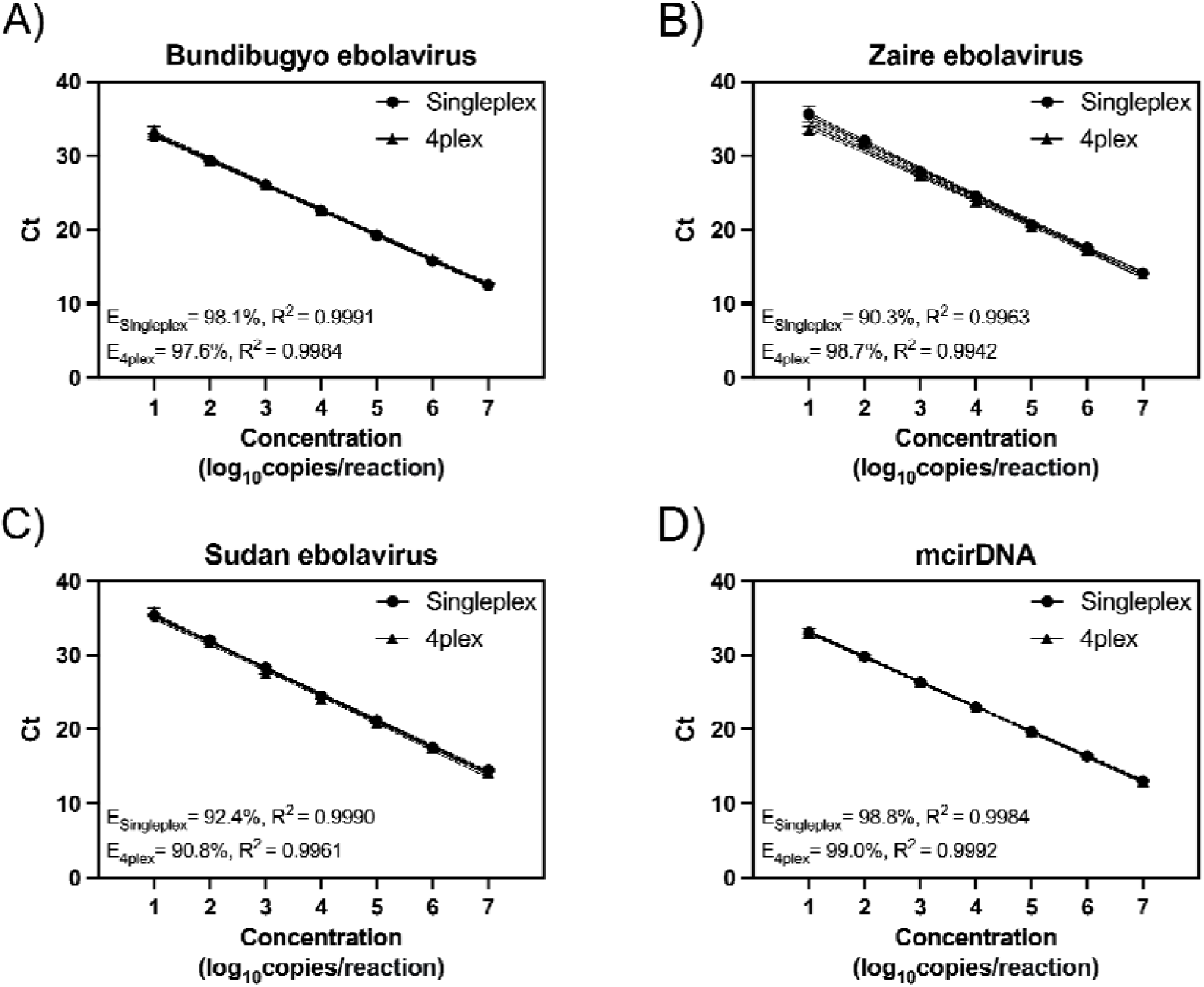
Standard curves for Bundibugyo ebolavirus (BDBV), Zaire ebolavirus (Z-EBOV), Sudan ebolavirus (S-EBOV), and mitochondrial circular DNA (mcirDNA) TaqMan RT-qPCR assays in singleplex and four-target multiplex formats. We generated standard curves for A) BDBV, B) Z-EBOV, C) S-EBOV, and D) the mcirDNA internal control assays using serial dilutions of synthetic RNA or DNA gene fragments. Data points show the mean and standard deviation of triplicate reactions. Solid lines indicate simple linear regressions, and dotted lines indicate the corresponding 95% confidence intervals. E indicates the RT-qPCR amplification efficiency.

**Table 2:** Abridged Limit of detection (LOD) analysis for Bundibugyo ebolavirus (BDBV), Zaire ebolavirus (Z-EBOV), Sudan ebolavirus (S-EBOV), and mitochondrial circular DNA (mcirDNA) across TaqMan singleplex, duplex, four-target multiplex and SYBR Green RT-qPCR assay formats. We determined the 95% limit of detection (LOD_95_) using synthetic RNA or DNA gene fragments quantified by digital PCR. We defined the LOD_95_ as the lowest concentration detected in at least 95% of replicate reactions (n=21). Only results at the concentration meeting the LOD_95_ criterion are shown; complete data are in Table A1.

| Target | Assay Type | Assay | Concentration (copies/reaction) | Detection (%) | Ct Mean | %CV |
| --- | --- | --- | --- | --- | --- | --- |
| **BDBV** | TaqMan | Singleplex | 5 | 95.2 | 34.1 | 3.4 |
|  |  | Duplex | 5 | 100 | 34.3 | 3.0 |
|  |  | Multiplex | 5 | 100 | 34.0 | 2.3 |
|  | SYBR Green | Singleplex | 50 | 100 | 29.3 | 1.1 |
| **mcirDNA** | TaqMan | Duplex | 5 | 100 | 33.2 | 2.7 |
|  |  | Multiplex | 5 | 100 | 34.7 | 2.4 |
| **Z-EBOV** | TaqMan | Multiplex | 5 | 95.2 | 34.9 | 4.4 |
| **S-EBOV** |  | Multiplex | 5 | 100 | 33.3 | 2.4 |

In contrived clinical samples, each assay detected its intended viral target across serial dilutions (Figure 3; Figure A4; R^2^ ≥ 0.99). We observed no cross-reactivity among the three tested ebolavirus species under the conditions tested. The mcirDNA internal control produced a consistent cycle threshold (Ct) across whole viral RNA concentrations, as expected from the constant human plasma background.

**Figure 3:**
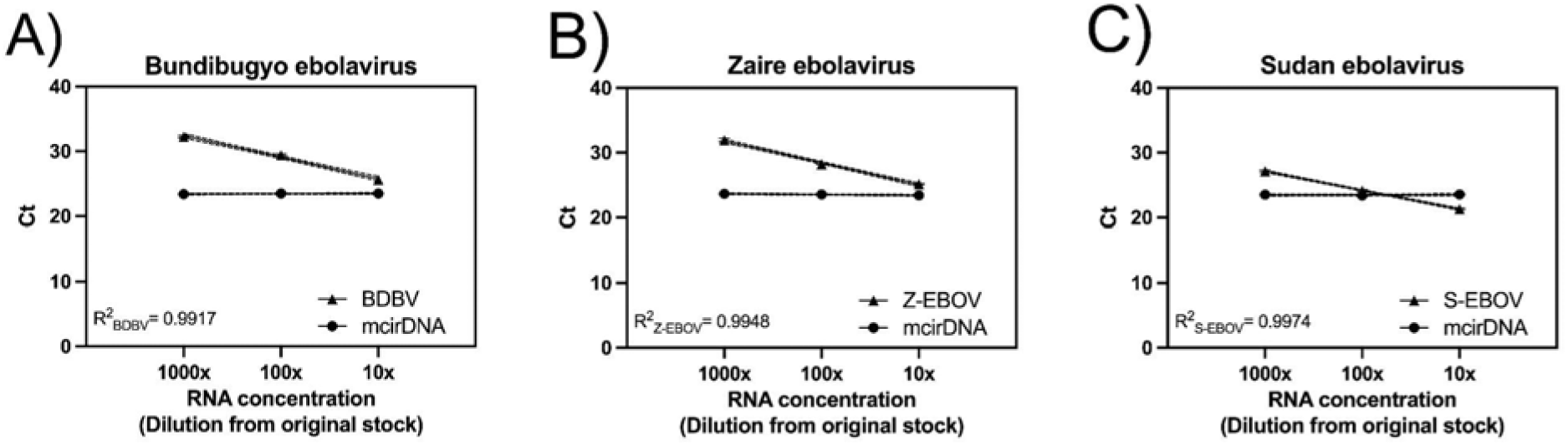
Detection of Bundibugyo ebolavirus (BDBV), Zaire ebolavirus (Z-EBOV), and Sudan ebolavirus (S-EBOV) whole viral RNA in contrived clinical samples using the four-target multiplex assay. We generated contrived clinical samples by spiking whole viral RNA from each ebolavirus species into a healthy human plasma background at three concentrations prepared by 10-fold serial dilutions of stock RNA. Each assay showed linear detection of its intended target across the dilution series, with no cross-reactivity among the other ebolavirus species. The mitochondrial circular DNA (mcirDNA) internal control maintained a consistent cycle threshold (Ct) value across each viral RNA concentration, reflecting the constant human plasma background. Data points show the mean and standard deviation of triplicate reactions. Solid lines indicate simple linear regressions, and dotted lines indicate the corresponding 95% confidence intervals.

### Performance of the probe-free SYBR Green assay

We evaluated the BDBV assay in a singleplex SYBR Green RT-qPCR format to provide a probe-free alternative for settings where TaqMan probe synthesis or supply is constrained. We selected final forward and reverse primer concentrations of 150nM based on amplification efficiency between 90–110%, linearity, and observed sensitivity across the tested conditions (Figure A5). The SYBR Green assay achieved an amplification efficiency of 99.6%, with an R^2^ value ≥ 0.99, and an LOD_95_ of 50 copies per reaction (Figure 4; Table 2). The assay also detected BDBV whole viral RNA in contrived clinical samples and showed no cross-reactivity with Z-EBOV or S-EBOV under the conditions tested. Although the probe-free format preserved specificity, its LOD_95_ was 10-fold higher that that of theTaqMan assays, indicating reduced analytical sensitivity.

**Figure 4:**
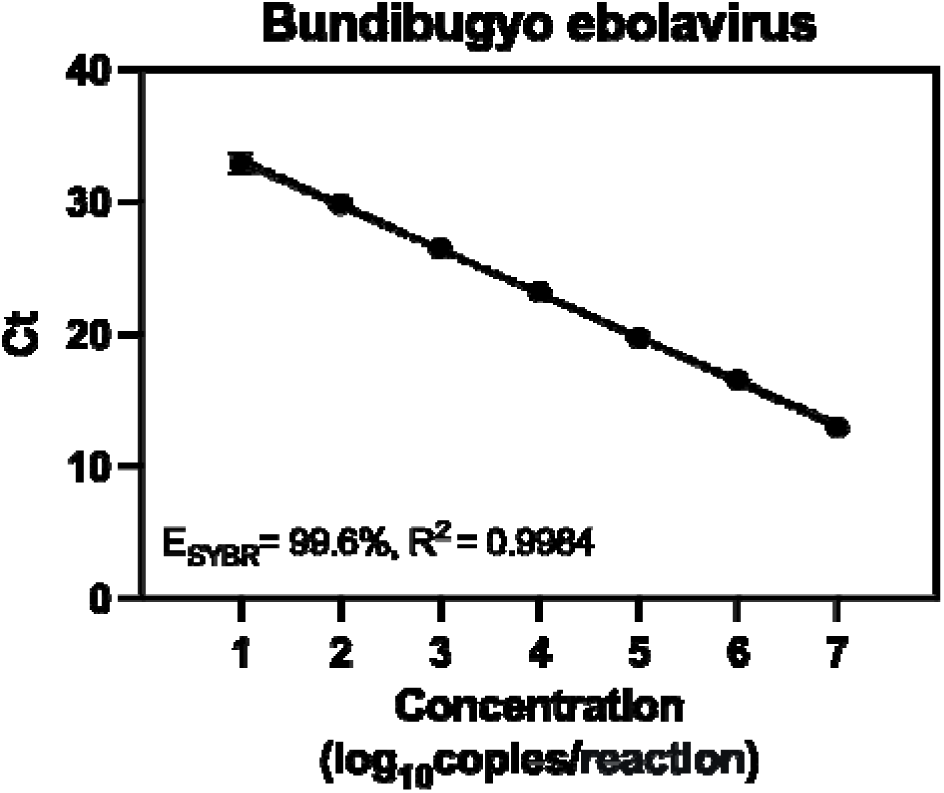
Standard curve for the Bundibugyo ebolavirus (BDBV) singleplex SYBR Green RT-qPCR assay. We generated the standard curve using serial dilutions of synthetic BDBV RNA gene fragments. Data points show the mean and standard deviation of triplicate reactions. The solid line indicates a simple linear regression, and dotted lines indicate the corresponding 95% confidence interval. E denotes RT-qPCR amplification efficiency.

### Rapid detection of Bundibuygo ebolavirus using TaqMan RT-qPCR on a portable qPCR instrument

We evaluated whether the BDBV singleplex assay and the BDBV-mcirDNA duplex assay could be adapted to a portable, rapid-cycling workflow. Using the Mic qPCR cycler, we shortened the reverse transcription, denaturation, and annealing steps, while benchmarking performance against the standard QuantStudio 6 Flex workflow (Table 3, Table A2). The optimized Mic protocol reduced total runtime from 65 to 35 minutes without compromising assay performance or analytical sensitivity.

**Table 3:**
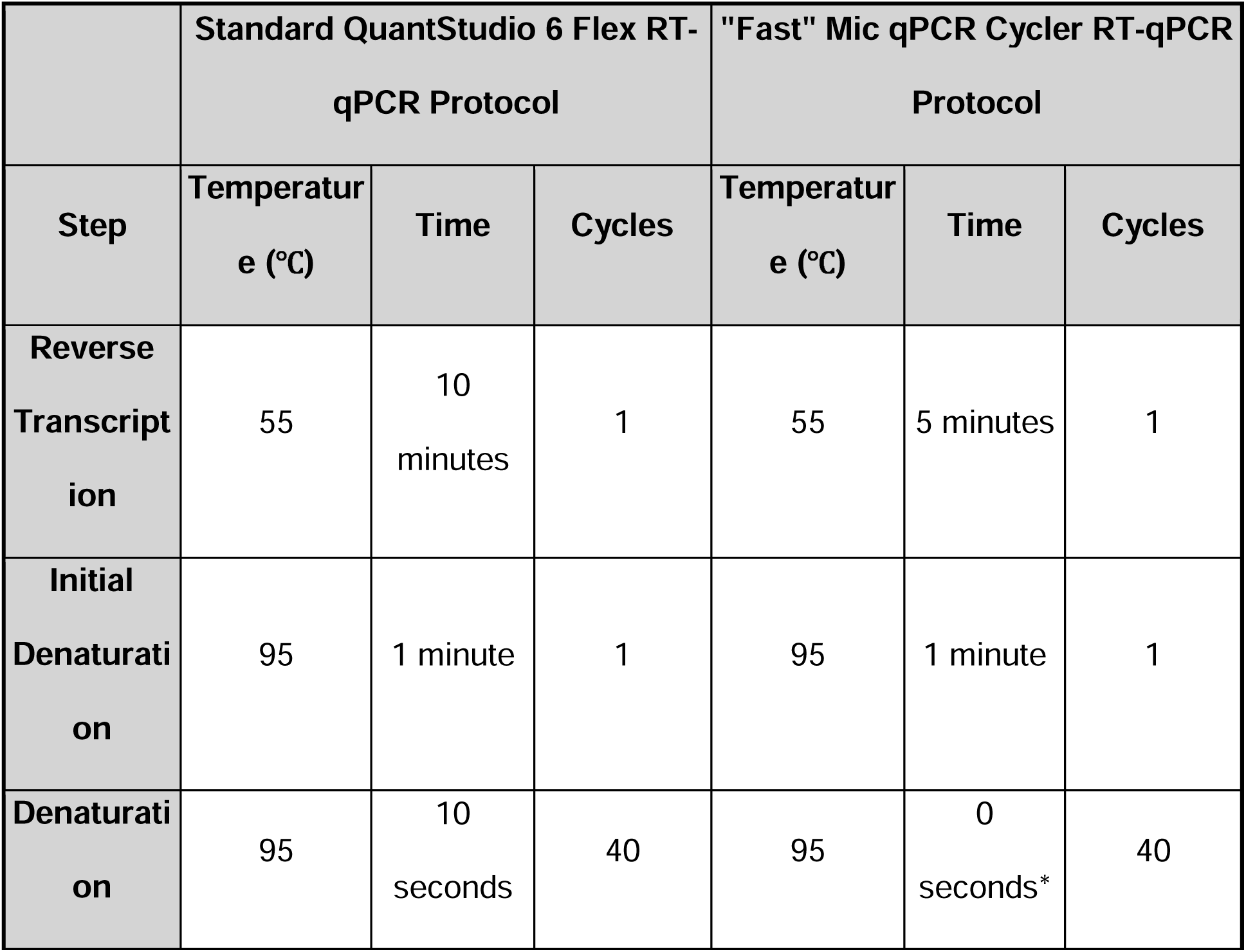
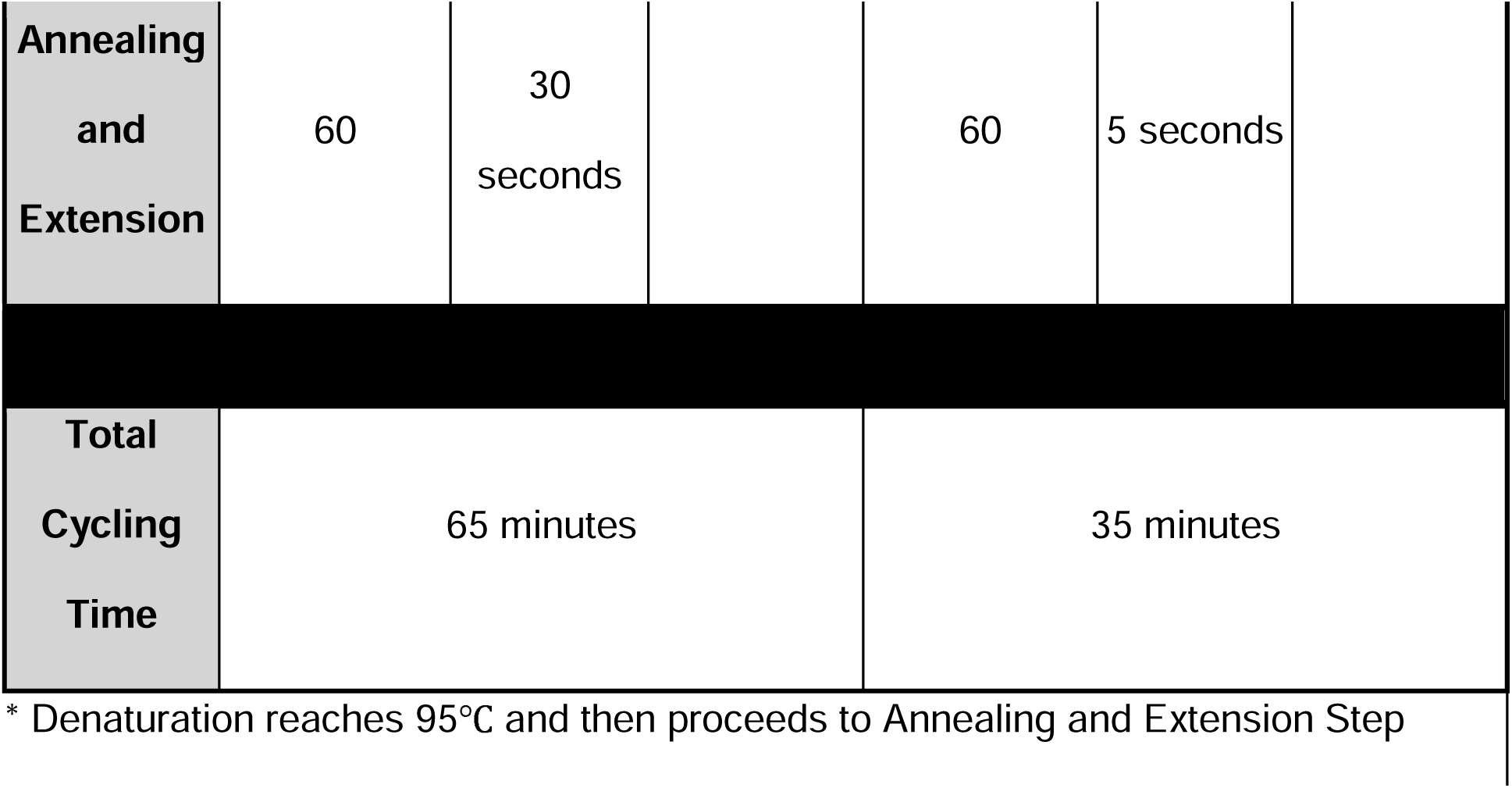
Standard and rapid thermocycling protocols for TaqMan RT-qPCR assays on the QuantStudio 6 Flex and Mic qPCR cycler. The standard QuantStudio 6 Flex protocol is shown at left, and the modified Mic qPCR cycler protocol is shown at right. Shortening the reverse-transcription, denaturation, and annealing steps reduced total runtime from 65 to 35 minutes while preserving assay performance and limit of detection.

We evaluated assay performance on the Mic qPCR cycler using synthetic DNA or RNA gene fragments at known concentrations and contrived clinical samples. Under the rapid-cycling protocol, the singleplex and duplex formats achieved amplification efficiencies of 97.1–105.6%, compared with 98.3–98.8% under standard cycling conditions on the QuantStudio 6 Flex, and all standard curves had R^2^ values ≥ 0.99 (Figures 1 and 5). Both formats maintained an LOD_95_ of 5 copies per reaction, matching the sensitivity of the corresponding QuantStudio 6 Flex workflows despite the shortened thermocycling profile (Table A3). The assays also showed linear detection of whole viral BDBV RNA across serial dilutions in contrived clinical samples (R^2^ ≥ 0.97; Figure A6). Neither assay detected whole viral RNA from Z-EBOV or S-EBOV under the conditions tested. In the duplex format, the mcirDNA internal control maintained a consistent Ct across BDBV concentrations, as expected from the constant human plasma background.

**Figure 5:**
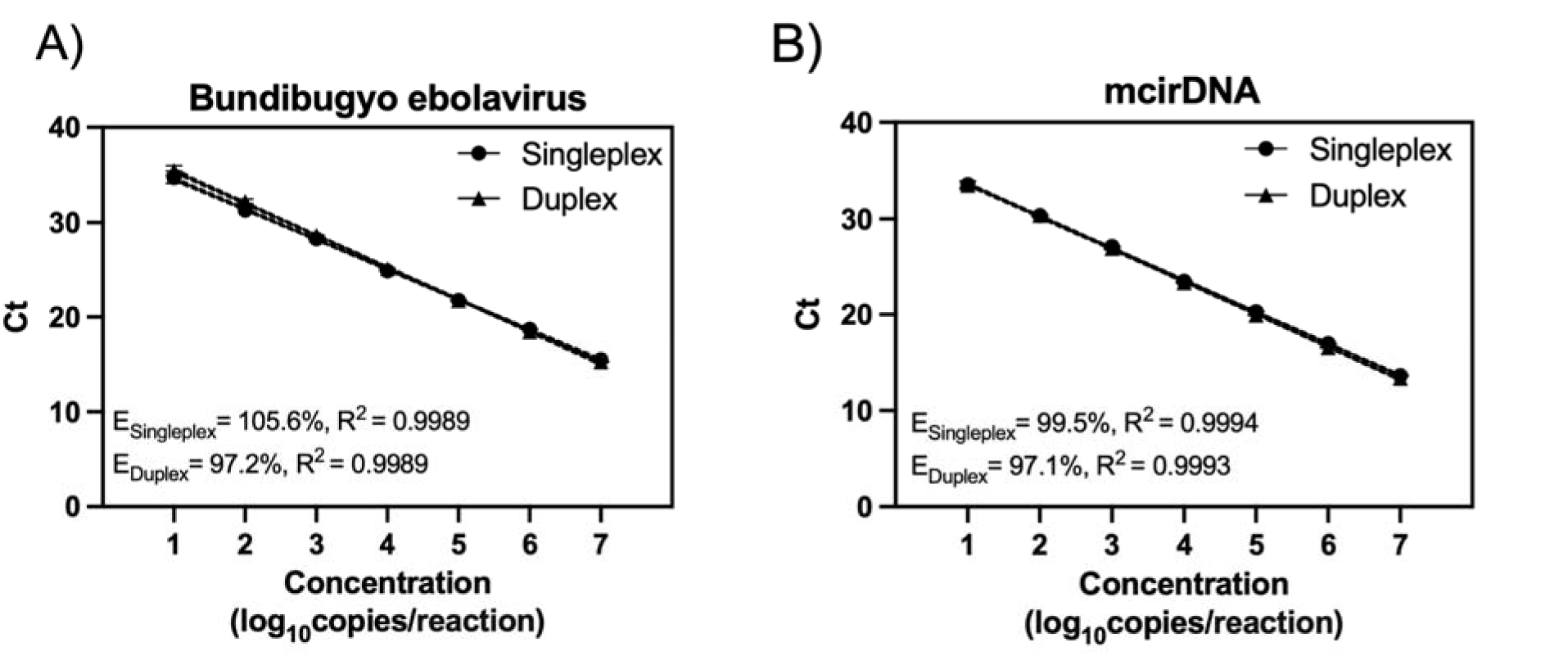
Standard curves for Bundibugyo ebolavirus (BDBV) and mitochondrial circular DNA (mcirDNA) TaqMan RT-qPCR assays in singleplex and duplex formats on the Mic qPCR cycler under rapid-cycling conditions. We generated standard curves for A) BDBV and B) mcirDNA internal control using serial dilutions of synthetic RNA or DNA gene fragments with the modified Mic qPCR protocol. Data points show the mean and standard deviation of triplicate reactions. Solid lines indicate simple linear regressions, and dotted lines indicate the corresponding 95% confidence interval. E denotes the RT-qPCR amplification efficiency.

### Performance with lyophilized RT-qPCR reagents

We evaluated a lyophilized RT-qPCR mastermix (LyoPrime Luna® Probe One-Step RT-qPCR Mix) on the QuantStudio 6 Flex and Mic qPCR cycler to reduce cold-chain requirements for BDBV testing in decentralized settings. We tested the BDBV assay in singleplex and duplex formats and benchmarked performance against the liquid Luna Probe One-Step RT-qPCR Kit. The lyophilized formulation preserved assay performance, linearity, and LODs across both instruments.

Using synthetic DNA or RNA gene fragments, the lyophilized assay achieved amplification efficiencies of 101.3-107.5% in singleplex format, and 96.9-99.9% in duplex format across both instruments (Figure 6). All standard curves had R^2^ values ≥ 0.99. Both assay formats maintained an LOD_95_ of 5 copies per reaction for each target on the QuantStudio 6 Flex and the Mic qPCR cycler (Table 4).

**Figure 6:**
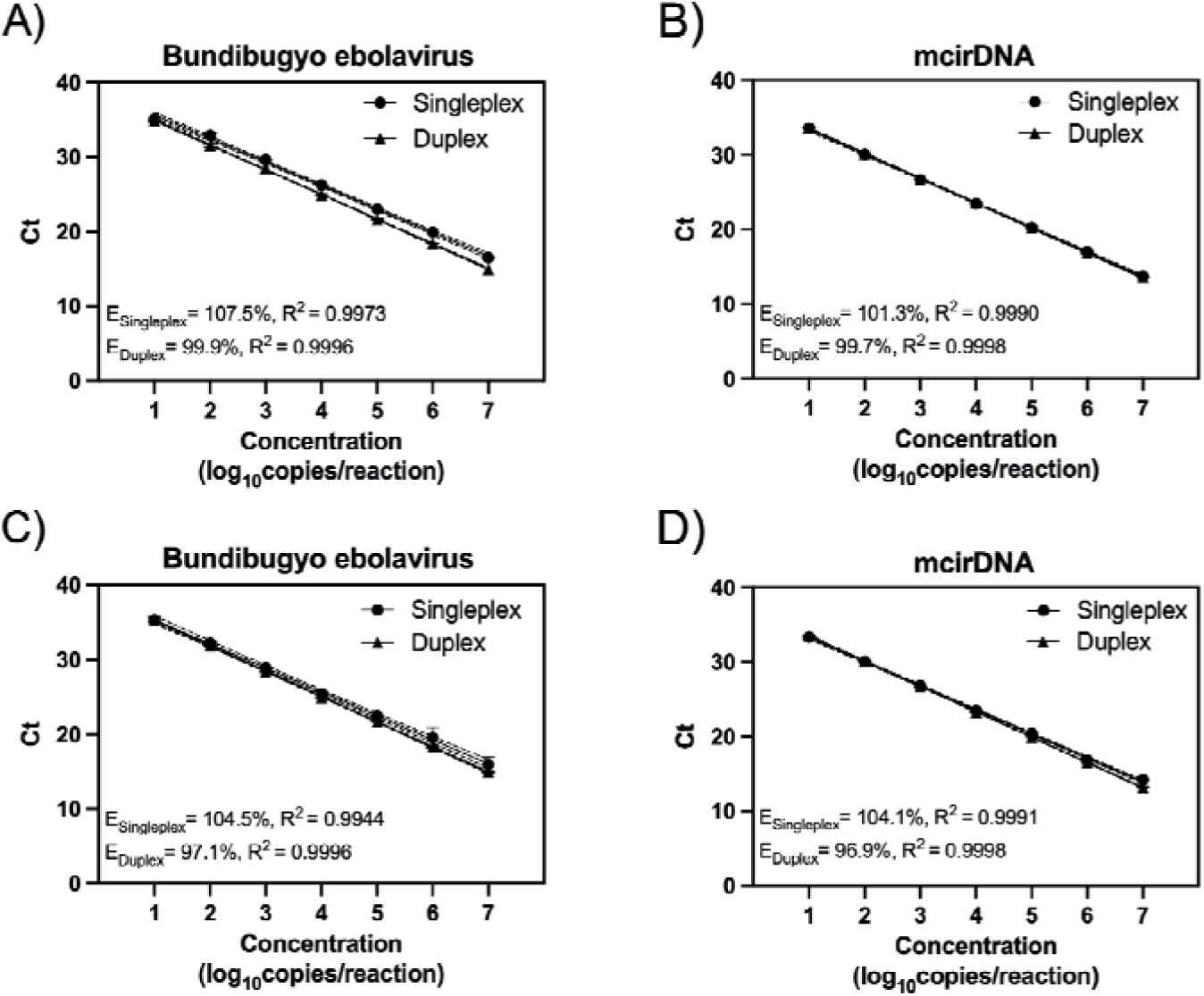
Standard curves for Bundibugyo ebolavirus (BDBV) and mitochondrial circular DNA (mcirDNA) TaqMan RT-qPCR assays in singleplex and duplex formats using lyophilized RT-qPCR mastermix on the QuantStudio 6 Flex and Mic qPCR cycler. We generated standard curves for A) BDBV on QuantStudio 6 Flex, B) the mcirDNA internal control on QuantStudio 6 Flex, C) BDBV on Mic qPCR cycler, and D) mcirDNA on the Mic qPCR cycler using serial dilutions of synthetic RNA gene fragments with lyophilized RT-qPCR mastermix. Data points show the mean and standard deviation of duplicate reactions. Solid lines indicate simple linear regressions, and dotted lines indicate the corresponding 95% confidence intervals. E denotes the RT-qPCR standard curve efficiency.

**Table 4:**
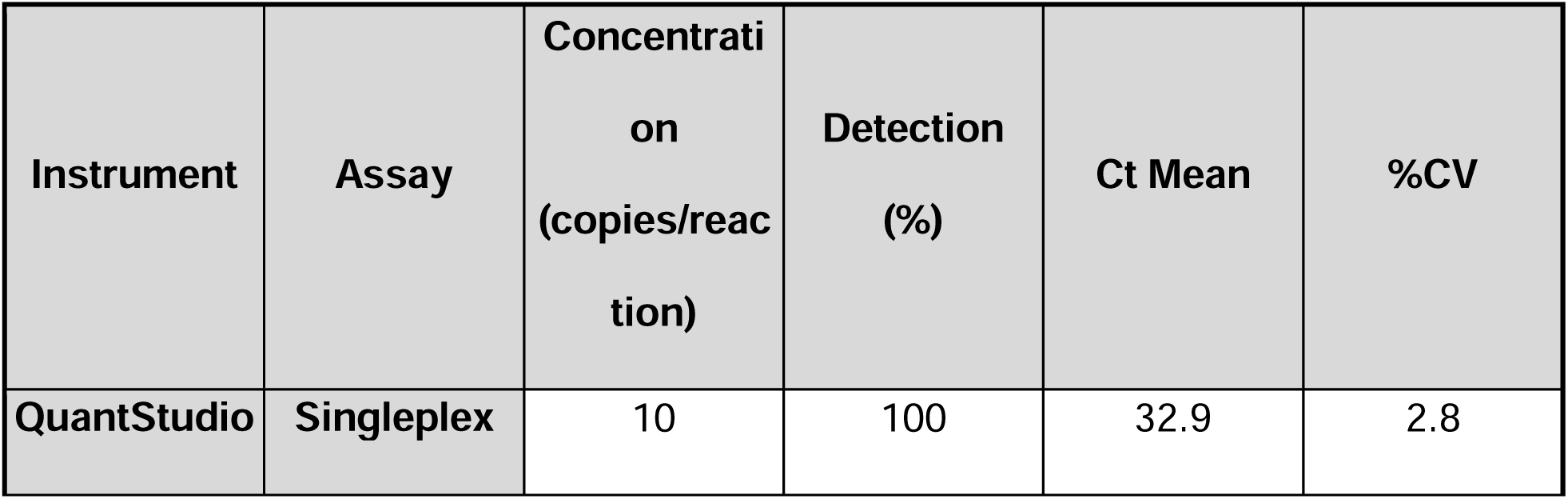
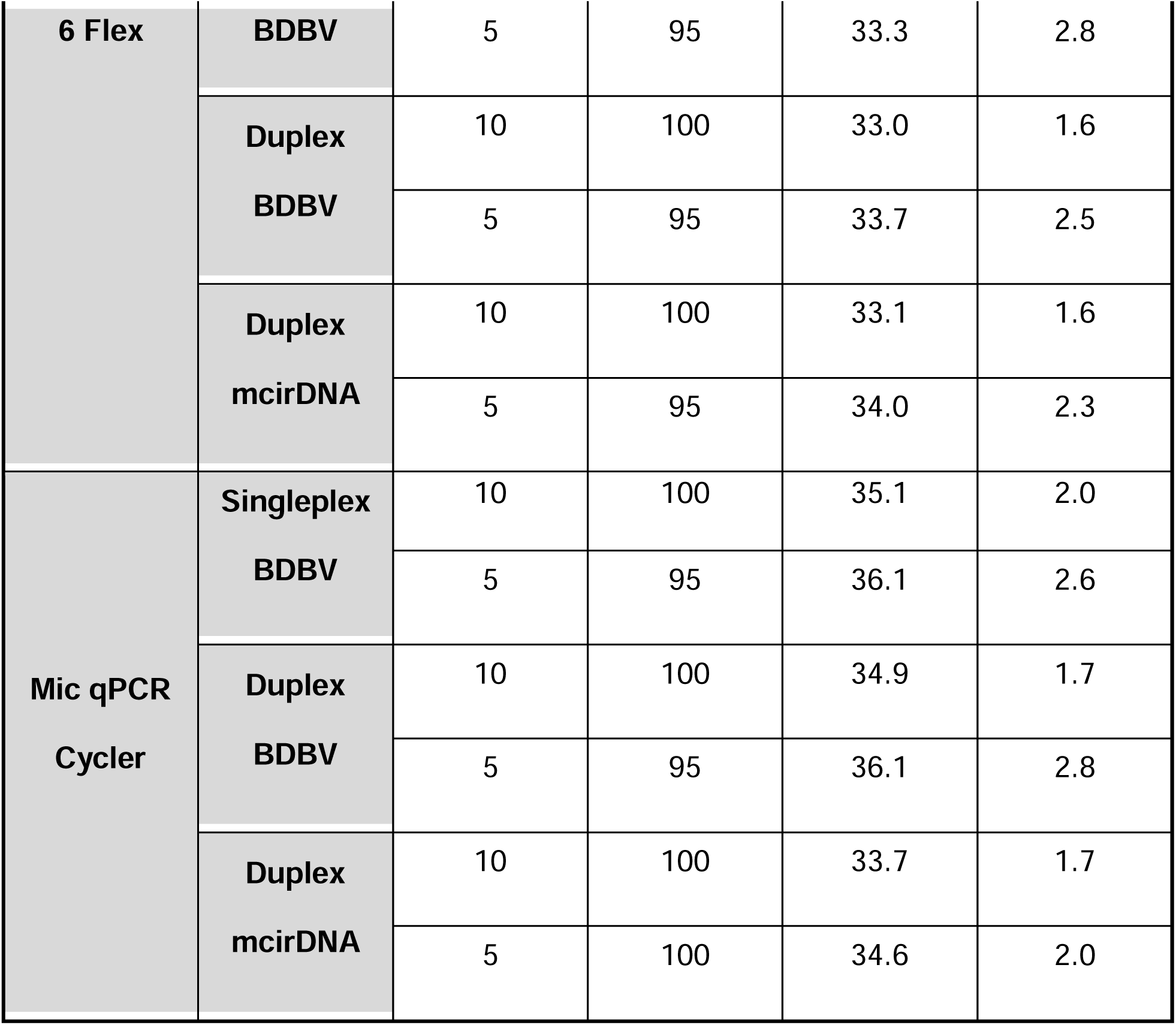
Limit of detection (LOD) analysis for Bundibugyo ebolavirus (BDBV) and mitochondrial circular DNA (mcirDNA) in singleplex and duplex RT-qPCR assay formats using lyophilized RT-qPCR mastermix on the QuantStudio 6 Flex and the Mic qPCR cycler. We determined the 95% limit of detection (LOD_95_) using synthetic RNA or DNA gene fragments quantified by digital PCR and amplified with lyophilized RT-qPCR mastermix on both instruments. We defined the LOD_95_ as the lowest concentration detected in at least 95% of replicate reactions (n=20)

Using contrived clinical samples, the lyophilized assays showed linear detection of whole viral BDBV RNA across serial dilutions (Figure A7, R^2^ ≥ 0.99). Neither assay format detected whole viral RNA from Z-EBOV or S-EBOV under the conditions tested. In the duplex format, the mcirDNA internal control maintained a consistent Ct across BDBV concentrations, as expected from the constant human plasma background.

### Specificity in human plasma background

To determine whether human-derived material interfered with assay performance or produced false-positive signals, we tested 20 replicates of pooled healthy human plasma background using the BDBV singleplex, duplex, and four-target multiplex TaqMan assays and the BDBV SYBR Green assay. No off-target amplification occurred in any assay format. In the duplex and multiplex assays, the mcirDNA internal control produced consistent Ct values across all replicates, as expected from the normalized human plasma background.

## Discussion

We developed, optimized, and analytically validated RT-qPCR assays that expand open, non-proprietary options for sensitive and specific detection of BDBV, Z-EBOV, and S-EBOV. These assays address a critical detection and surveillance gap highlighted by the current BDBV outbreak in the DRC and Uganda. By implementing the assays in singleplex, duplex, multiplex, and probe-free formats, we provide complementary options that laboratories can adapt to different testing needs, outbreak-response, and broader pathogen surveillance.

The portable workflow extends these assays beyond conventional centralized laboratory settings, which is critical for effective outbreak response (*16*). We adapted the BDBV singleplex and duplex TaqMan assays to the Mic qPCR cycler and validated lyophilized reagents to reduce cold-chain requirements. The Mic qPCR cycler has performed well in other near point of care (POC) settings, including suitcase-based malaria testing, and prior studies have shown that modified cycling conditions can substantially shorten qPCR run times while preserving assay efficiency (*17–19*). In our study, the portable workflow maintained analytical sensitivity and linearity while reducing runtime from 65 to 35 minutes. These features could expand access to rapid BDBV testing in decentralized laboratories and mobile outbreak-response settings. Because the workflow still requires conventional nucleic acid extraction, however, it should be considered portable rather than fully POC. Future integration with simplified sample-processing methods could further reduce equipment and infrastructure requirements.

Existing BDBV diagnostics include commercial assays and several published laboratory-developed tests, but important gaps remain in transparency, analytical validation, and deployability (*20–27*). Proprietary primer and probe sequences can limit independent evaluation against newly generated outbreak genomes and make it harder for laboratories to assess or adapt assay performance as the virus evolves. Commercial dependence may also create supply and cold-chain constraints during outbreaks. Published noncommercial assays help address these limitations, but some rely on degenerate primers, show mismatches to circulating outbreak sequences, or do not report the full analytical data needed to evaluate sensitivity, specificity, and robustness for outbreak surveillance (*21–27*). By contrast, our BDBV assay showed complete sequence identity across the historical and 2026 outbreak genomes evaluated and maintained an LOD_95_ of 5 copies per reaction across singleplex, duplex, and multiplex formats, two instruments, rapid and standard cycling conditions, and liquid and lyophilized reagents. These features provide laboratories with a transparent, analytically characterized, and adaptable complement to existing diagnostic options.

Several limitations should guide future development, validation and implementation. First, we evaluated assay specificity using contrived clinical samples containing BDBV, Z-EBOV, and S-EBOV whole viral RNA, but broader testing against other filoviruses, including Marburg virus, would strengthen the specificity assessment. Although NCBI BLAST analysis predicted no off-target amplification against other pathogens, empirical testing across a wider panel remains important. Second, the current workflows require conventional nucleic acid extraction, limiting use to settings with appropriate laboratory infrastructure and biosafety capacity. Integrating simplified, field-compatible sample processing could move the assays closer to true POC use. Third, we were unable to test specimens from patients with confirmed BDBV infection, so clinical validation is still needed to establish real-world sensitivity and specificity. Continued *in silico* monitoring will also be important as additional BDBV genomes become available to ensure that primer and probe binding sites remain conserved.

Disease outbreaks require detection methods that can be developed and adapted quickly, rigorously evaluated, and made freely available to support surveillance and response. We developed these assays as flexible tools for BDBV detection, outbreak response, and future research, while reducing dependence on limited commercial options during periods of high demand. We shared the assay designs, validation data, and protocols as we developed them in real time through Ampliphi (https://www.ampliphi.bio), a new open-access platform for rapidly disseminating diagnostic assays and supporting their adoption. Beyond the current BDBV outbreak, this approach to transparent assay development, analytical validation, and rapid dissemination could provide a framework for responding to future emerging pathogens before, during, and between outbreaks.

## Methods

### Molecular assay design

Bundibugyo virus (BDBV) genomes (n=26) were downloaded from NCBI Virus with filters applied (Tax ID: 3052458, Nucleotide Completeness: complete, access date: May 17, 2026). These genomes were aligned in Geneious using MAFFT, and several TaqMan qPCR primer and probe combinations were designed using Geneious combined with Primer3. Once outbreak specific sequences were published, genomes were obtained from Pathoplexis (n=16 on June 4, 2026). The chosen BDBV design had 100% sequence alignment to all historical BDBV sequences as well as to the newly released outbreak sequences. Previously designed Zaire ebolavirus (Z-EBOV) and Sudan ebolavirus (S-EBOV) assays were utilized to facilitate rapid multiplexing (ongoing work, unpublished). Z-EBOV (n=135) and S-EBOV (n=121) genomes were downloaded from NCBI Virus (from collection date: 01/01/2022, access date: 02/06/2025, min length: 18000), aligned in Geneious Prime, and specific primers and TaqMan probes were designed to detect each species. In addition, a previously published human internal control assay targeting circulating human mitochondrial DNA in human blood (mcirDNA, Meddeb et al., 2019) was adapted to a TaqMan qPCR assay and previously optimized (ongoing work, unpublished work). This assay was included as a human internal control for plasma samples.

### *In silico* analysis of designs

Primers and probes were mapped to on-target and off-target sequences in Geneious Prime to test specificity *in silico*. The sequence sets used for analysis were the same as used for design, with all assays mapped against the three different ebolavirus species. The BDBV assay was additionally tested against sequences as they became available during the outbreak (n=533 with complete sequencing data in the amplicon region as of August 10th, 2026). An assay was predicted to detect a sequence if both primers and probe mapped with ≤ 3 mismatches. In addition, all primer and probe sequences were evaluated for broader cross-reactivity using NCBI BLAST.

### Samples and controls

Synthetic double stranded DNA gene fragments (Twist Bioscience, sequences provided in Table A4) targeting specific, unique regions of Z-EBOV (Viral Protein 24), S-EBOV (L segment, RNA-dependent RNA polymerase), BDBV (L segment, RNA-dependent RNA polymerase), and mcirDNA (Cytochrome C Oxidase Subunit III) were utilized as synthetic standards. Viral targets (BDBV, Z-EBOV, and S-EBOV) underwent *in vitro* transcription (IVT) and DNase treatment using the HiScribe® T7 High Yield RNA Synthesis Kit (New England Biolabs, E2040L), followed by purification using RNAClean XP beads (Beckman Colter, A63987). All synthetic standards were then quantified using the Qubit™ RNA High Sensitivity (HS) Kit (Invitrogen, Q32852) for viral targets or the Qubit™ DNA High Sensitivity (HS) Kit (Invitrogen, Q33231) for mcirDNA. Based on calculated quantification, the synthetic standards were diluted and normalized to 1E8 copies/ L aliquots. Use of all kits followed instructions as recommended by the manufacturer

For contrived sample experimentation, whole viral RNA (Z-EBOV and S-EBOV provided by NEIDL, Boston University; BDBV obtained from BEI) was spiked into pooled extracted human plasma sample matrix obtained from healthy, non-infected individuals (Innovative Research, IPLAWBK2E50ML). Briefly, human plasma was extracted using the Quick-DNA/RNA MagBead Extraction Kit (Zymo Research, R2131) on the KingFisher™ Flex Magnetic Particle Processor (ThermoFisher) with the 96 Deep-Well Head as recommended by the manufacturer. The extract was pooled to create a standard and consistent human background signal in subsequent RT-qPCR experimentation.

### TaqMan qPCR assay evaluation and validation

Initial Taqman qPCR evaluation was performed on the QuantStudio 6 Flex utilizing the Luna Probe One-Step RT-qPCR Kit (No ROX) (NEB, E3007E) as 10 L reactions in triplicate following manufacturer’s recommendations (see protocols for reaction composition and cycling conditions). All analytical evaluations were performed on synthetic gene fragment RNA or DNA on a standard curve ranging from 1E7–1E1 copies/reaction. Assay performance was then evaluated based on qPCR standard curve efficiency (ranging from 90–110%), linearity, and observed sensitivity of the assay based on imputed material concentration. A sample was considered positive if at least 2/3 sample triplicates amplified. Amplified replicates were then averaged and plotted as the mean Ct with error bars.

All assay designs underwent initial evaluation as singleplex FAM TaqMan qPCR assays at three different primer-probe concentrations (primer concentrations/probe concentration): 200nM/200nM, 400nM/200nM, and 600nM/200nM Probe (Integrated DNA Technologies). After selecting optimal primer-probe concentrations for each singleplex FAM TaqMan probe assay, a duplex assay (composed of BDBV (FAM) and mcirDNA (HEX)) and a multiplex assay (composed of BDBV (FAM), Z-EBOV (Cy5), S-EBOV (ROX), and mcirDNA (HEX)) were evaluated in singleplex, duplex, or multiplex form simultaneously.

### SYBR Green BDBV assay optimization and evaluation

A singleplex SYBR Green BDBV assay was optimized utilizing the following primer concentrations (forward and reverse primers at equimolar concentrations): 100nM, 150nM, 200nM, and 450nM. All SYBR Green experiments were conducted on a QuantStudio 6 Flex following methodology described above. Evaluation and performance of this assay was conducted using the Power SYBR™ Green RNA-to-CT™ 1-Step Kit (Applied Biosystems™, 4389986) as 10 L reactions in triplicate following manufacturer’s recommendations (see protocol for reaction composition and cycling conditions).

### Adaptation of TaqMan assays to the Mic qPCR Cycler

We adapted our BVBD singleplex and duplex assays to run on the Mic qPCR cycler (Bio Molecular Systems), a portable, fast cycling compatible qPCR machine. We initially ran the thermocycling profile of the assay as originally run on the QuantStudio 6 Flex, and systematically changed the cycling conditions to assess performance with the goal of shortening the overall run time. The conditions tested were: 1) original profile, 2) shortening the denaturation hold time to 0 seconds, 3) shortening the annealing hold time to 10 seconds, and 4) shortening the reverse transcription step to 5 min and the annealing hold time to 5 seconds. After optimization, we ran all remaining experiments with the run profile of (4).

All analytical evaluations were performed on synthetic gene fragment RNA or DNA on a standard curve ranging from 1E7–1E1 copies/reaction. Assay performance was then evaluated based on qPCR standard curve efficiency (ranging from 90–110%), linearity, and observed sensitivity of the assay based on imputed material concentration.

### Evaluation of a lyophilized RT-qPCR mastermix

We evaluated our BDBV TaqMan singleplex and duplex assays on a lyophilized RT-qPCR mastermix (LyoPrime Luna® Probe One-Step RT-qPCR Mix with UDG, NEB, L4001P) to ensure compatibility. Each well of the lyophilized mastermix was rehydrated with 18 L of water containing primers and probes to give final assay concentrations of 400nM primer and 200nM probe in a 20 L reaction. Next, 2 L of sample was added to each well and duplicate 10 L reactions were run on the Mic qPCR cycler or QuantStudio 6 Flex. A sample was considered positive if both sample duplicates amplified. Amplified replicates were then averaged and plotted with the mean Ct with error bars.

### Limit of detection determination

To determine the limit of detection (LOD) of our optimized assays on the QuantStudio 6 Flex using the standard qPCR cycling conditions, we ran replicates (n=21 for non-lyophilized mastermix or n=20 for lyophilized mastermix) of the following synthetic gene fragment concentrations for the BDBV singleplex TaqMan assay, duplex assay, multiplex assay, and SYBR assay: 5E1 (non-lyophilized mastermix only), 1E1, and 5E0 copies/reaction. Similarly, we determined the LOD of the assays on the Mic qPCR cycler by running replicates (n=21 for non-lyophilized mastermix or n=20 for lyophilized mastermix) of the following synthetic gene fragment concentrations for the BDBV TaqMan singleplex and duplex: 1E1 and 5E0 copies/reaction.

The gene fragments were accurately quantified and normalized on digital PCR (dPCR) using the QIAcuity OneStep Advanced Probe Kit (Qiagen, 250131) following the manufacturer’s recommendations. The LOD_95_ was defined as the concentration where at least 20/21 (for non-lyophilized) or 19/20 (for lyophilized mastermix) replicates amplified.

### Assay specificity evaluation against human sample matrix

We ran replicates (n=20) of extracted pooled healthy human plasma background on the BDBV assays on the QuantStudio 6 Flex under standard conditions to evaluate assay specificity against a human sample matrix. Specificity was defined here as the Negative Percent Agreement (NPA), calculated as the percentage of known negative samples correctly identified as negative.

### Statistical Analysis

All data generated via RT-qPCR was analyzed and graphed in GraphPad Prism (version 10.5.0). Standard curves were calculated using standard linear regression to obtain 95% confidence intervals, assess goodness of fit (R^2^ values), and to establish qPCR efficiencies (via slope value).

## Data Availability

All data produced in the present study are available upon reasonable request to the authors

https://www.ampliphi.bio/t/quantitative-pcr-assays-for-sensitive-and-specific-detection-of-bundibugyo-ebolavirus/15

## Acknowledgements

The following reagent was obtained through BEI Resources, NIAID, NIH:

- RNA from Bundibugyo ebolavirus, Prototype Isolate #811250 (200706291 Uganda), NR-31812

We would like to thank the NEIDL at Boston University for providing RNA from Zaire ebolavirus and Sudan ebolavirus.

The use of AI was used to streamline language, support sentence structure and flow, and assist in citation formatting. AI was not utilized for data, figure, or table generation.

---

**Figure A5:**
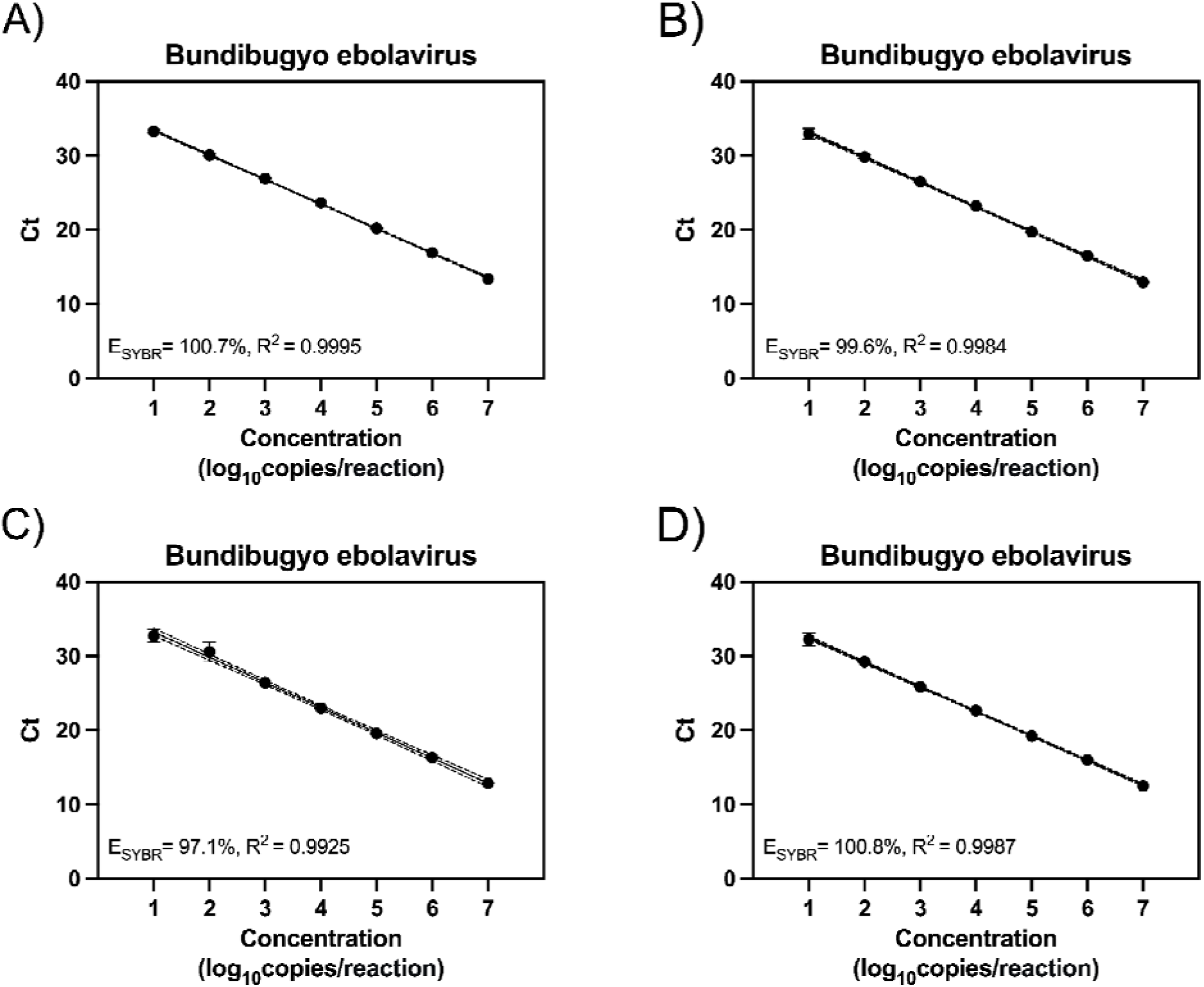
Optimization of primer concentration for the Bundibugyo ebolavirus qPCR SYBR Green singleplex assay. We evaluated the SYBR Green singleplex assay at the following forward and reverse primer concentrations: A) 100nM, B) 150nM, C) 200nM, D) 450nM. Standard curves were generated with synthetic RNA gene fragments. Data points represent the mean and standard deviation of triplicates. The solid line represents a simple linear regression and the dotted lines represent the 95% confidence interval of the linear regression.

**Figure A6:**
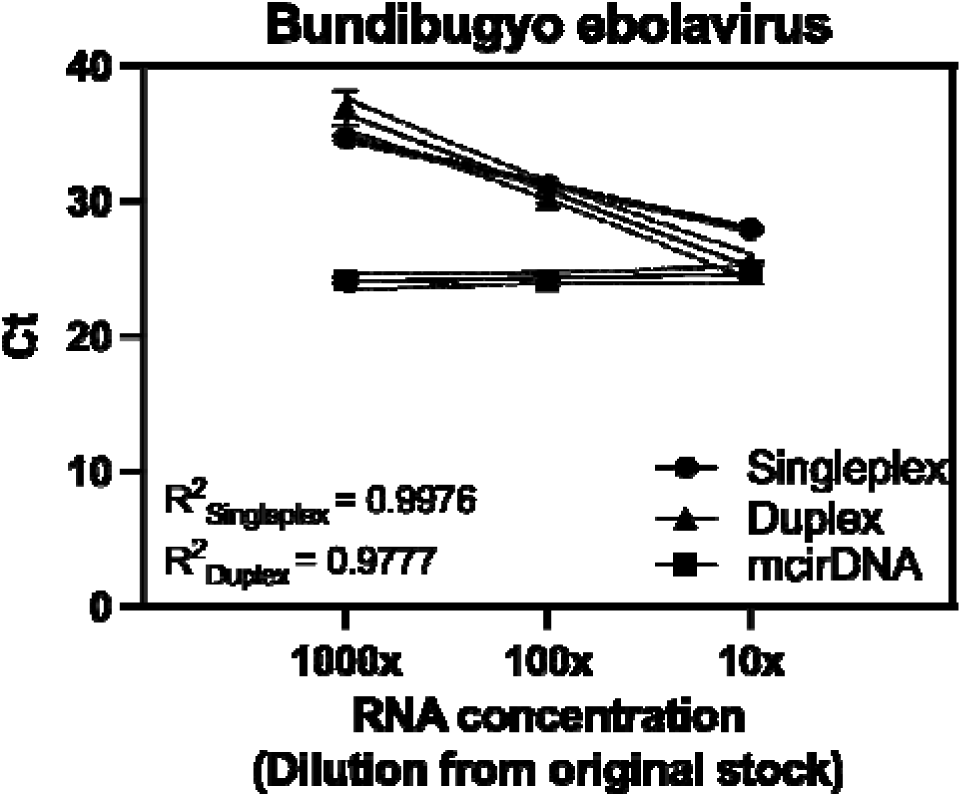
Detection of Bundibugyo ebolavirus (BDBV) whole viral RNA in contrived clinical samples utilizing the fast Mic qPCR cycler protocol. We generated contrived clinical samples by spiking BDBV whole viral RNA into normalized healthy human plasma background at three concentrations prepared by 10-fold serial dilutions of stock RNA. The BDBV assay showed linear detection across the dilution series in both singleplex and duplex formats. For the duplex assay, the mitochondrial circular DNA (mcirDNA) internal control showed a consistent cycle threshold (Ct) across BDBV dilutions, reflecting the constant human plasma background. Data points show the mean and standard deviation of triplicate reactions. Solid lines show simple linear regressions, and dotted lines show the 95% confidence intervals for the regressions.

**Figure A7:**
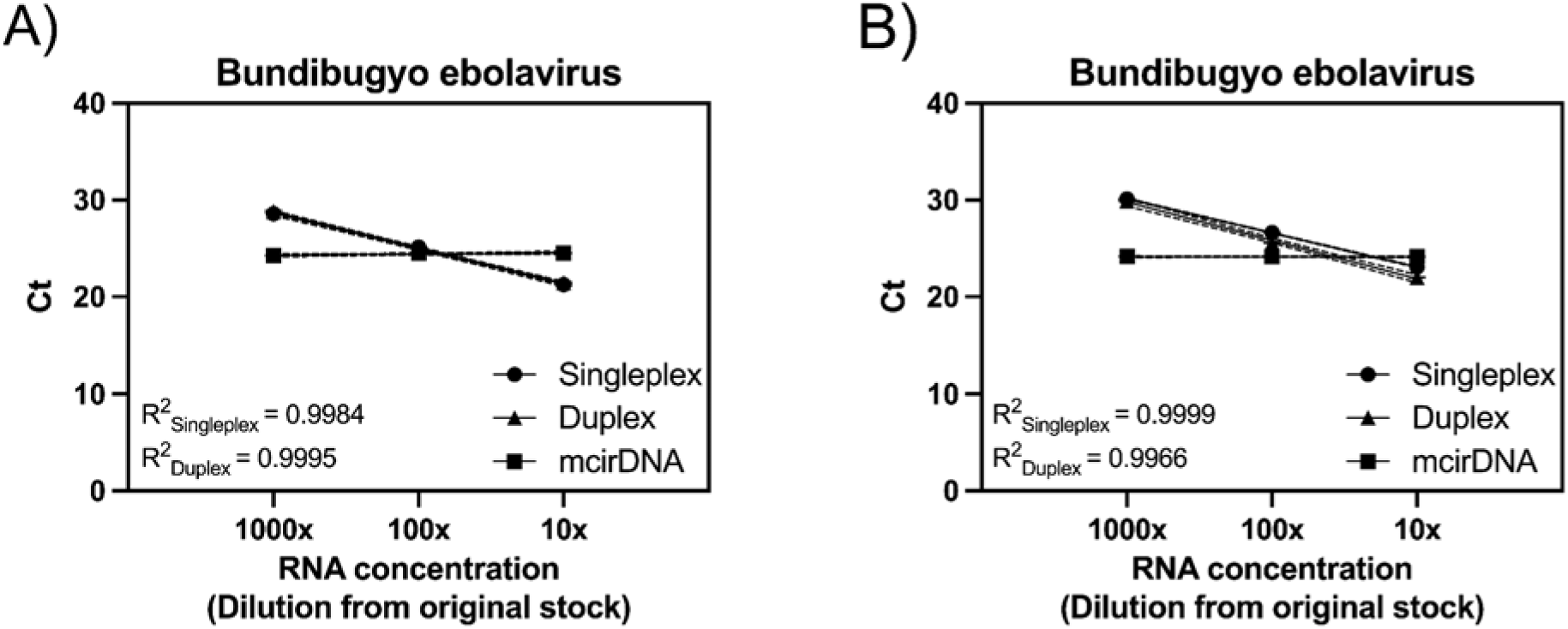
Detection of Bundibugyo ebolavirus (BDBV) whole viral RNA in contrived clinical samples utilizing lyophilized mastermix reagents on the QuantStudio 6 and the Mic qPCR cycler. We generated contrived clinical samples by spiking BDBV whole viral RNA into normalized healthy human plasma background at three concentrations prepared by 10-fold serial dilutions of stock RNA which were run on lyophilized RT-qPCR mastermix on both the a) QuantStudio 6 Flex and the b) Mic qPCR cycler. The BDBV assay showed linear detection across the dilution series in both singleplex and duplex formats on both instruments. For the duplex assay, the mitochondrial circular DNA (mcirDNA) internal control showed a consistent cycle threshold (Ct) across BDBV dilutions, reflecting the constant human plasma background on both instruments. Data points show the mean and standard deviation of duplicate reactions. Solid lines show simple linear regressions, and dotted lines show the 95% confidence intervals for the regressions.

## Citations

1. World Health Organization. Ebola disease caused by Bundibugyo virus, Democratic Republic of the Congo & Uganda. https://www.who.int/emergencies/disease-outbreak-news/item/2026-DON602.

2. DRC Ministry of Public Health, Hygiene and Social Welfare. Official declaration Ebola outbreak, 15 May 2026. Logistics Cluster Website. 2026 May 15. https://logcluster.org/en/documents/drc-ministry-public-health-hygiene-and-social-welfare-official-declaration-ebola-outbreak.

3. World Health Organization. Epidemic of Ebola Disease caused by Bundibugyo virus in the Democratic Republic of the Congo and Uganda determined a public health emergency of international concern. 2026 May 17. https://www.who.int/news/item/17-05-2026-epidemic-of-ebola-disease-in-the-democratic-republic-of-the-congo-and-uganda-determined-a-public-health-emergency-of-international-concern.

4. Centers for Disease Control and Prevention. Ebola Outbreak: Current Situation. 2026. https://www.cdc.gov/ebola/situation-summary/index.html.

5. Cheng N, Ye RZ, Li YY, Kargbo KB, Ren LL, Cao WC. Global distribution and genetic diversity of orthoebolaviruses: Mapping and evolutionary analysis. One Health. 2025 Dec;21:101236. doi: 10.1016/j.onehlt.2025.101236. PMID: 41141938; PMCID: PMC12547875.

6. Jain S, Martynova E, Rizvanov A, Khaiboullina S, Baranwal M. Structural and Functional Aspects of Ebola Virus Proteins. Pathogens. 2021 Oct 15;10(10):1330. doi: 10.3390/pathogens10101330. PMID: 34684279; PMCID: PMC8538763.

7. Centers for Disease Control and Prevention. Ebola Disease Basics. 2026. https://www.cdc.gov/ebola/about/index.html.

8. Centers for Disease Control and Prevention. History of Ebola Outbreaks. 2026. https://www.cdc.gov/ebola/outbreaks/index.html.

9. Bettini A, Lapa D, Garbuglia AR. Diagnostics of Ebola virus. Front Public Health. 2023;11:1123024. doi: 10.3389/fpubh.2023.1123024. PMID: 36908455; PMCID: PMC9995846.

10. World Health Organization. Diagnostic testing for Ebola and Marburg virus diseases: interim guidance, 20 December 2024. World Health Organization; 2025. https://iris.who.int/handle/10665/380073. doi: 10.2471/B09221.

11. Sealy TK. Laboratory Response to Ebola — West Africa and United States. MMWR Suppl. 2016;65. doi: 10.15585/mmwr.su6503a7.

12. Mérens A, Bigaillon C, Delaune D. Ebola virus disease: Biological and diagnostic evolution from 2014 to 2017. Médecine Mal Infect. 2018 Mar 1;48(2):83–94. doi: 10.1016/j.medmal.2017.11.002.

13. World Health Organization, Regional Office for Africa. Bundibugyo Ebola virus | Continental preparedness and response plan: June-November 2026. 2026 Jun. https://www.afro.who.int/publications/bundibugyo-ebola-virus-continental-preparedness-and-response-plan-june-november-2026.

14. Bio Molecular Systems. Choosing a Real-Time PCR Cycler. 2021 Dec 21. https://biomolecularsystems.com/choosing-a-real-time-pcr-cycler-that-works-for-your-lab/.

15. Meddeb R, Dache ZAA, Thezenas S, Otandault A, Tanos R, Pastor B, et al. Quantifying circulating cell-free DNA in humans. Sci Rep. 2019 Mar 26;9(1):5220. doi: 10.1038/s41598-019-41593-4. PMID: 30914716; PMCID: PMC6435718.

16. Matthews Q, da Silva SJR, Norouzi M, Pena LJ, Pardee K. Adaptive, diverse and de-centralized diagnostics are key to the future of outbreak response. BMC Biol. 2020 Oct 28;18(1):153. doi: 10.1186/s12915-020-00891-4.

17. Carlier L, Baker SC, Huwe T, Yewhalaw D, Haileselassie W, Koepfli C. qPCR in a suitcase for rapid Plasmodium falciparum and Plasmodium vivax surveillance in Ethiopia. PLOS Glob Public Health. 2022 Jul 27;2(7):e0000454. doi: 10.1371/journal.pgph.0000454. PMID: 36962431; PMCID: PMC10021179.

18. Bustin SA. How to speed up the polymerase chain reaction. Biomol Detect Quantif. 2017 Jun 20;12:10–4. doi: 10.1016/j.bdq.2017.05.002. PMID: 28702368; PMCID: PMC5496742.

19. Bustin SA, Kirvell S, Nolan T, Shipley GL. FlashPCR: Revolutionising qPCR by Accelerating Amplification through Low ΔT Protocols. Int J Mol Sci. 2024 Feb 28;25(5):2773. doi: 10.3390/ijms25052773. PMID: 38474020; PMCID: PMC10932470.

20. Roche Diagnostics. Roche rapidly develops a PCR test for Ebola within six days. 2026. https://diagnostics.roche.com/global/en/news-listing/2026/roche-rapidly-develops-a-pcr-test-for-ebola-within-six-days-as-r.html.

21. Woolsey C, Borisevich V, Agans KN, O’Toole R, Fenton KA, Harrison MB, et al. A Highly Attenuated Panfilovirus VesiculoVax Vaccine Rapidly Protects Nonhuman Primates Against Marburg Virus and 3 Species of Ebola Virus. J Infect Dis. 2023 Nov 15;228(Suppl 7):S660–70. doi: 10.1093/infdis/jiad157. PMID: 37171813; PMCID: PMC11009496.

22. O’Donnell KL, Haase JA, Henderson CW, Gathright BR, Fletcher P, Rhoderick JF, et al. A Single-Dose Bundibugyo Virus Vaccine Protects Macaques Within 3 Days. bioRxiv. 2026 Jun 14. https://www.biorxiv.org/content/10.64898/2026.06.14.732188v1. doi: 10.64898/2026.06.14.732188.

23. Kozak R, He S, Kroeker A, de La Vega MA, Audet J, Wong G, et al. Ferrets Infected with Bundibugyo Virus or Ebola Virus Recapitulate Important Aspects of Human Filovirus Disease. J Virol. 2016 Sep 29;90(20):9209–23. doi: 10.1128/jvi.01033-16.

24. Towner JS, Sealy TK, Khristova ML, Albariño CG, Conlan S, Reeder SA, et al. Newly Discovered Ebola Virus Associated with Hemorrhagic Fever Outbreak in Uganda. PLOS Pathog. 2008 Nov 21;4(11):e1000212. doi: 10.1371/journal.ppat.1000212.

25. Falzarano D, Feldmann F, Grolla A, Leung A, Ebihara H, Strong JE, et al. Single immunization with a monovalent vesicular stomatitis virus-based vaccine protects nonhuman primates against heterologous challenge with Bundibugyo ebolavirus. J Infect Dis. 2011 Nov;204 Suppl 3(Suppl 3):S1082–1089. doi: 10.1093/infdis/jir350. PMID: 21987745; PMCID: PMC3189995.

26. Robert Koch Institute. Ebolavirus-Diagnostik: Nachweis von Bundibugyo-Virus [Ebolavirus diagnostics: Detection of Bundibugyo virus]. 2026 Jun 12. https://www.rki.de/DE/Themen/Infektionskrankheiten/Infektionskrankheiten-A-Z/E/Ebola/Diagnostik-Protokolle.html.

27. Liu J, Ochieng C, Wiersma S, Ströher U, Towner JS, Whitmer S, et al. Development of a TaqMan Array Card for Acute-Febrile-Illness Outbreak Investigation and Surveillance of Emerging Pathogens, Including Ebola Virus. J Clin Microbiol. 2016 Jan;54(1):1–13. doi: 10.1128/jcm.02257-15.

