## Supplemental Tables and Figures for "Rapid and portable reverse-transcription quantitative PCR assays for Bundibugyo ebolavirus detection"

Author affiliations: Broad Institute of MIT and Harvard, Cambridge, Massachusetts, USA (K. McMahon, S. Nielsen, H. Knoll, R. Talwar, D. Thompson, C. Wilkason, A. Ozonoff, E. Stachler, P. C. Sabeti); Boston Children’s Hospital, Boston, Massachusetts, USA (A. Ozonoff); Harvard Medical School, Boston, Massachusetts, USA (A. Ozonoff); Harvard University, Cambridge, Massachusetts, USA (P. C. Sabeti); Howard Hughes Medical Institute, Chevy Chase, Maryland, USA (P. C. Sabeti)

^†^These senior authors contributed equally to this article

*Corresponding author

Elyse Stachler

415 Main St., Cambridge, MA 02142, USA

617-820-7518

Table A1: Limit of detection (LOD) analysis for Bundibugyo ebolavirus (BDBV), Zaire ebolavirus (Z-EBOV), Sudan ebolavirus (S-EBOV), and mitochondrial circular DNA (mcirDNA) in the singleplex, duplex, and multiplex (4-plex) RT-qPCR assay formats. We evaluated the 95% limit of detection (LOD_95_) for the singleplex TaqMan, duplex TaqMan, multiplex TaqMan and SYBR Green BDBV assays formats using synthetic RNA gene fragments quantified by digital PCR. We defined the LOD_95_ as the lowest concentration that produced amplification in at least 95% of replicate reactions (n=21).

| **Target** | **Assay Type** | **Assay** | **Concentration**  **(copies/reaction)** | **Detection (%)** | **Ct Mean** | **%CV** |
| --- | --- | --- | --- | --- | --- | --- |
| **BDBV** | TaqMan | Singleplex | 50 | 100 | 30.6 | 0.9 |
|  |  |  | 10 | 100 | 33.3 | 1.6 |
|  |  |  | 5 | 95.2 | 34.1 | 3.4 |
|  |  | Duplex | 50 | 100 | 30.6 | 1.2 |
|  |  |  | 10 | 100 | 33.0 | 1.8 |
|  |  |  | 5 | 100 | 34.3 | 3.0 |
|  |  | Multiplex | 50 | 100 | 30.6 | 0.9 |
|  |  |  | 10 | 100 | 32.8 | 1.8 |
|  |  |  | 5 | 100 | 34.0 | 2.3 |
|  | SYBR Green | Singleplex | 50 | 100 | 29.3 | 1.1 |
|  |  |  | 10 | 85.7 | 32.6 | 2.4 |
|  |  |  | 5 | 57.1 | 33.1 | 2.6 |
| **mcirDNA** | TaqMan | Duplex | 50 | 100 | 29.7 | 1.2 |
|  |  |  | 10 | 100 | 31.9 | 1.6 |
|  |  |  | 5 | 100 | 33.2 | 2.7 |
|  |  | Multiplex | 50 | 100 | 31.0 | 0.8 |
|  |  |  | 10 | 100 | 33.4 | 2 |
|  |  |  | 5 | 100 | 34.7 | 2.4 |
| **Z-EBOV** | TaqMan | Multiplex | 50 | 100 | 31.3 | 1.2 |
|  |  |  | 10 | 100 | 34.0 | 2.5 |
|  |  |  | 5 | 95.2 | 34.9 | 4.4 |
| **S-EBOV** | TaqMan | Multiplex | 50 | 100 | 30.0 | 0.7 |
|  |  |  | 10 | 100 | 32.0 | 1.6 |
|  |  |  | 5 | 100 | 33.3 | 2.4 |

Table A2: Iterative fast cycling profiles evaluated on the Mic qPCR cycler. Version 1-4 of cycling profiles trialed on the Mic qPCR cycler on the Ebolavirus Bundibugyo (BDBV) singleplex TaqMan assay. qPCR cycling profiles were evaluated on a RNA standard curve ranging from 1E7–1E1 copies/reaction. Assay performance was then evaluated based on qPCR standard curve efficiency (ranging from 90–110%), linearity, and observed sensitivity of the assay based on imputed material concentration.

| **qPCR Protocol Version** | **qPCR Testing Conditions** | **Run Time** | **Concentrations Tested** | **Mean Ct** | **%CV** |
| --- | --- | --- | --- | --- | --- |
| **Version 1** | **Standard qPCR Parameters** | **62m** | 1.00E+07 | 16.4 | 0.31% |
|  |  |  | 1.00E+06 | 20.0 | 0.28% |
|  |  |  | 1.00E+05 | 22.7 | 0.16% |
|  |  |  | 1.00E+04 | 26.2 | 0.28% |
|  |  |  | 1.00E+03 | 29.4 | 0.56% |
|  |  |  | 1.00E+02 | 32.8 | 0.46% |
|  |  |  | 1.00E+01 | 36.9 | 2.82% |
| **Version 2** | **0s Denatuarization Step** | **56m** | 1.00E+07 | 16.6 | 0.09% |
|  |  |  | 1.00E+06 | 20.6 | 0.23% |
|  |  |  | 1.00E+05 | 23.1 | 0.20% |
|  |  |  | 1.00E+04 | 26.8 | 0.15% |
|  |  |  | 1.00E+03 | 29.8 | 0.64% |
|  |  |  | 1.00E+02 | 32.7 | 0.99% |
|  |  |  | 1.00E+01 | 36.0 | 2.61% |
| **Version 3** | **0s Denatuarization Step**  **10s Annealing Step** | **43m** | 1.00E+07 | 16.0 | 0.32% |
|  |  |  | 1.00E+06 | 20.1 | 0.26% |
|  |  |  | 1.00E+05 | 22.4 | 0.28% |
|  |  |  | 1.00E+04 | 26.3 | 0.29% |
|  |  |  | 1.00E+03 | 29.2 | 0.14% |
|  |  |  | 1.00E+02 | 32.3 | 0.79% |
|  |  |  | 1.00E+01 | 35.6 | 1.79% |
| **Version 4** | **0s Denatuarization Step**  **5s Annealing Step**  **5m RT Step** | **35m** | 1.00E+07 | 15.5 | 0.13% |
|  |  |  | 1.00E+06 | 18.7 | 0.17% |
|  |  |  | 1.00E+05 | 21.8 | 0.25% |
|  |  |  | 1.00E+04 | 24.9 | 0.14% |
|  |  |  | 1.00E+03 | 28.3 | 0.25% |
|  |  |  | 1.00E+02 | 31.3 | 0.06% |
|  |  |  | 1.00E+01 | 34.8 | 1.74% |

Table A3: Limit of detection (LOD) analysis for Bundibugyo ebolavirus (BDBV) and mitochondrial circular DNA (mcirDNA) in singleplex and duplex RT-qPCR assay formats on the Mic qPCR cycler under fast cycling conditions. We evaluated the 95% limit of detection (LOD_95_) for the singleplex and duplex TaqMan BDBV and mcirDNA assays using synthetic RNA or DNA gene fragments quantified by digital PCR. We defined the LOD_95_ as the lowest concentration that produced amplification in at least 95% of replicate reactions (n=21).

| **Assay** | **Concentration**  **(copies/reaction)** | **Detection (%)** | **Ct Mean** | **%CV** |
| --- | --- | --- | --- | --- |
| **Singleplex**  **BDBV** | 10 | 100 | 34.4 | 1.4 |
|  | 5 | 100 | 35.6 | 2.3 |
| **Duplex**  **BDBV** | 10 | 100 | 34.4 | 1.4 |
|  | 5 | 95.2 | 35.6 | 1.7 |
| **Duplex**  **mcirDNA** | 10 | 100 | 33.6 | 1.2 |
|  | 5 | 95.2 | 34.6 | 1.7 |

Table A4: Assay gene fragment sequences used as positive control sequences. Double stranded DNA gene fragments utilized as synthetic positive controls for the BDBV, Z-EBOV, S-EBOV, and mcirDNA assays.

| **Assay Name** | **Gene Fragment Sequences** |
| --- | --- |
| Bundibugyo ebolavirus | GAAATTAATACGACTCACTATAGGGTACCTAACCTACACGTCTACGCTTTCCTTGGATCTCACAAGGTACCGAGAGAATGAGTTAATTTATGATAACAATCCGTTAAAAGGTGGACTTAATTGCAACCTATCCTTTGATAATCCACTTTTCAAGGGCCAAAGGCTCAATATCATAGAGGAGGATTTGATTAGATTTCCTCATCTATCTGGGTGGGAACTTGCGAAAACCATCATTCAGTCCATTATCTCAGACAGCAATAACTCATCCACAGACCCCATTAGCAGTGGAGAAACACGATCATTCACAACTCACTTTCTCACATATCCTAAGGTTGGGCTCCTCTATAGTTTCGGCGCCATCGTGAGTTATTACTTAGGGAATACCATTATTAGGACCAAAAAGCTAGACCTCAGTCATTTTATGTATTACTTAACAACTCAAATCCATAATTTGCCACATCGCTCGTTGAGGATACTTAAGCCCACCTTTAAACATGTTAGTGTGATATCAAGACTAATGAGTAT |
| Zaire ebolavirus | GAAATTAATACGACTCACTATAGGGCAGCTGATTGACCAGTCTTTGATTGAGCCCTTAGCAGGAGCCCTTGGTCTGATCTCTGATTGGCTGCTAACAACCAACACTAACCATTTCAACATGCGAACACAACGTGTCAAGGAACAATTGAGCCTAAAAATGCTGTCGTTGATTCGATCCAATATTCTCAAGTTTATTAACAAATTGGATGCTCTACATGTCGTGAACTACAACGGATTGTTGAGCAGTATTGAAATTGGAACTCAAAATCATACAATCATCATAACTCGAACTAACATGGGTTTTCTGGTGGAGCTCCAAGAACCCGACAAATCGGCAATGAACCGCAAGAAGCCTGGGCCGGCGAAATTTTCCCTCCTTCATGAGTCTACACTGAAAGCATTTACACAAGGATCCTCGACACGAATGCAAAGTTTGATTCTTGAATTTAATAGCTCTCTTGCTATCTAACTAAGATGGAATACTTCATATTGAGCTAACTCATATATGCTGACTCAATAGTTATC |
| Sudan ebolavirus | GAAATTAATACGACTCACTATAGGGTTAGCAGCGGTGAAACAAGATCCTTCACAACCCACTTCTTAACGTATCCCAAAATAGGGCTCCTATACAGTTTTGGAGCCCTCATAAGTTTTTATTTGGGTAATACTATTCTGTGCACGAAAAAGATCGGACTCACAGAATTTCTATACTATCTCCAGAATCAGATCCACAACTTATCACACAGATCCCTTCGAATCTTCAAACCGACATTTAGACACTCAAGTGTCATGTCCAGGTTGATGGATATAGACCCCAACTTCTCAATATATATTGGTGGGACTGCAGGTGACCGTGGATTATCGGACGCTGCAAGATTATTTCTCCGAATTGCAATTTCAACTTTCTTGAGCTTTGTTGAGGAGTGGGTTATCTTTAGGAAGGCAAACATCCCACTATGGGTTATCTATCCTCTCGAAGGCCAACGCCCTGATCCTCCTGGCGAATTTTTGAACCGAGTAAAATCTCTAATTATTGGGACTGAAGATGATAAAAATAAAGGT |
| Mitochondrial circular DNA | TCTACACTTATCATCTTCACAATTCTAATTCTACTGACTATCCTAGAAATCGCTGTCGCCTTAATCCAAGCCTACGTTTTCACACTTCTAGTAAGCCTCTACCTGCACGACAACACATAATGACCCACCAATCACATGCCTATCATATAGTAAAACCCAGCCCATGACCCCTAACAGGGGCCCTCTCAGCCCTCCTAATGACCTCCGGTCTAGCCATGTGATTTCACTTCCACTCCATAACGCTCCTCATACTAGGCCTACTAACCAACACACTAACCATATACCAATGATGGCGCGATGTAACACGAGAAAGCACATACCAAGGCCACCACACACCACCTGTCCAAAAAGGCCTTCGATACGGGATAATCCTATTTATTACCTCAGAAGTTTTTTTCTTCGCAGGATTTTTCTGAGCCTTTTACCACTCCAGCCTAGCCCCTACCCCCCAACTAGGAGGGCACTGGCCCCCAACAGGCATCACCCCGCTAAATCCCCTA |

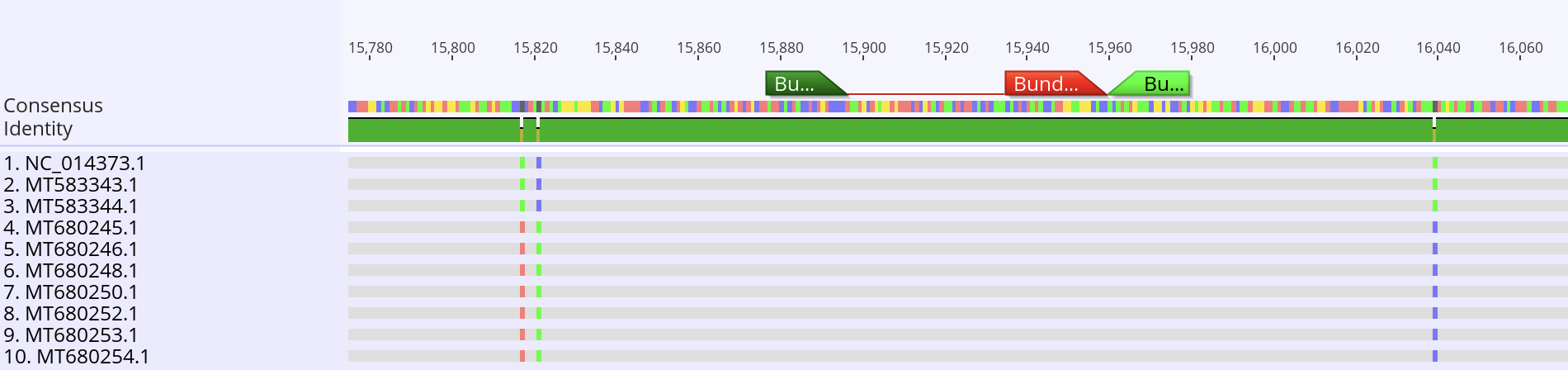

Figure A1: Alignment of the Bundibugyo ebolavirus (BDBV) RT-qPCR primers and probe to historic BVBV genomes. Complete historical BDBV genomes were aligned in Geneious; a representative view of the alignment is shown for readability. The locations of the forward primer, probe, and reverse primer are indicated at the top of the alignment. Although sequence variation is present at some positions in the surrounding genomic region, all three primer and probe binding sites show 100% sequence identity across the historical genomes.

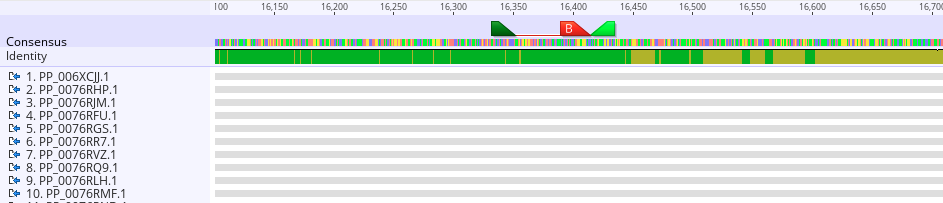

Figure A2: Alignment of the Bundibugyo ebolavirus (BDBV) RT-qPCR primers and probe to 2026 outbreak BVBV genomes. Complete BDBV genomes from the current 2026 outbreak (n=533 with complete sequencing data in the amplicon region as of August 10th, 2026) were aligned in Geneious; a representative view of the alignment is shown for readability. The locations of the forward primer, probe, and reverse primer are indicated at the top of the alignment. Although sequence variation is present at some positions in the surrounding genomic region, all three primer and probe binding sites show 100% sequence identity across the outbreak genomes.

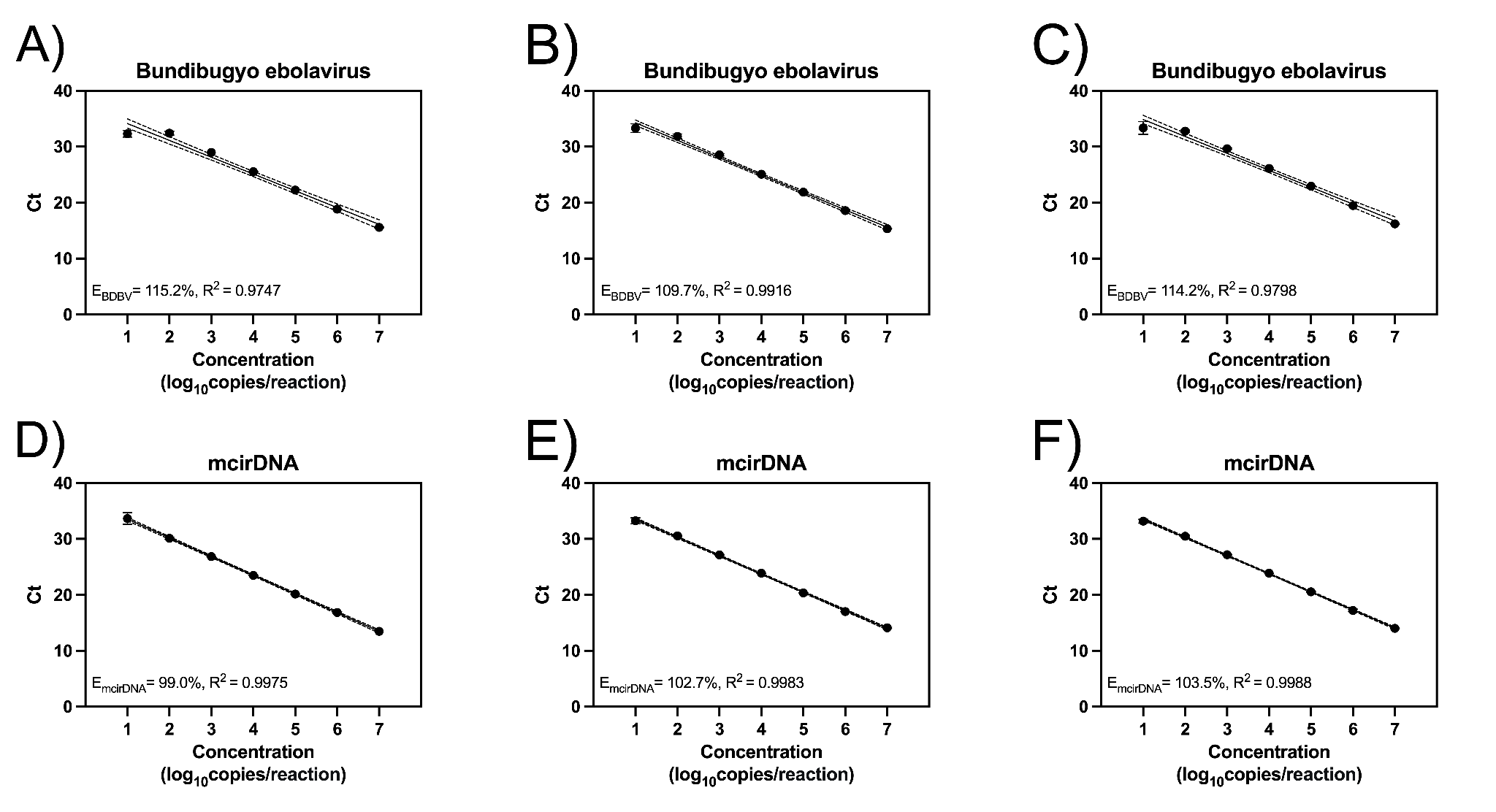

Figure A3: Optimization of primer and probe concentration for the Bundibugyo ebolavirus and mcirDNA qPCR TaqMan singleplex assays. We evaluated the Bundibugyo ebolavirus and mcirDNA qPCR TaqMan singleplex assays at the following primer/probe concentrations: A)/D) 200nM/200nM, B)/E) 400nM/200nM, and C)/F) 600nM/200nM. Standard curves were generated with synthetic RNA gene fragments. Data points represent the mean and standard deviation of triplicates. The solid line represents a simple linear regression and the dotted lines represent the 95% confidence interval of the linear regression.

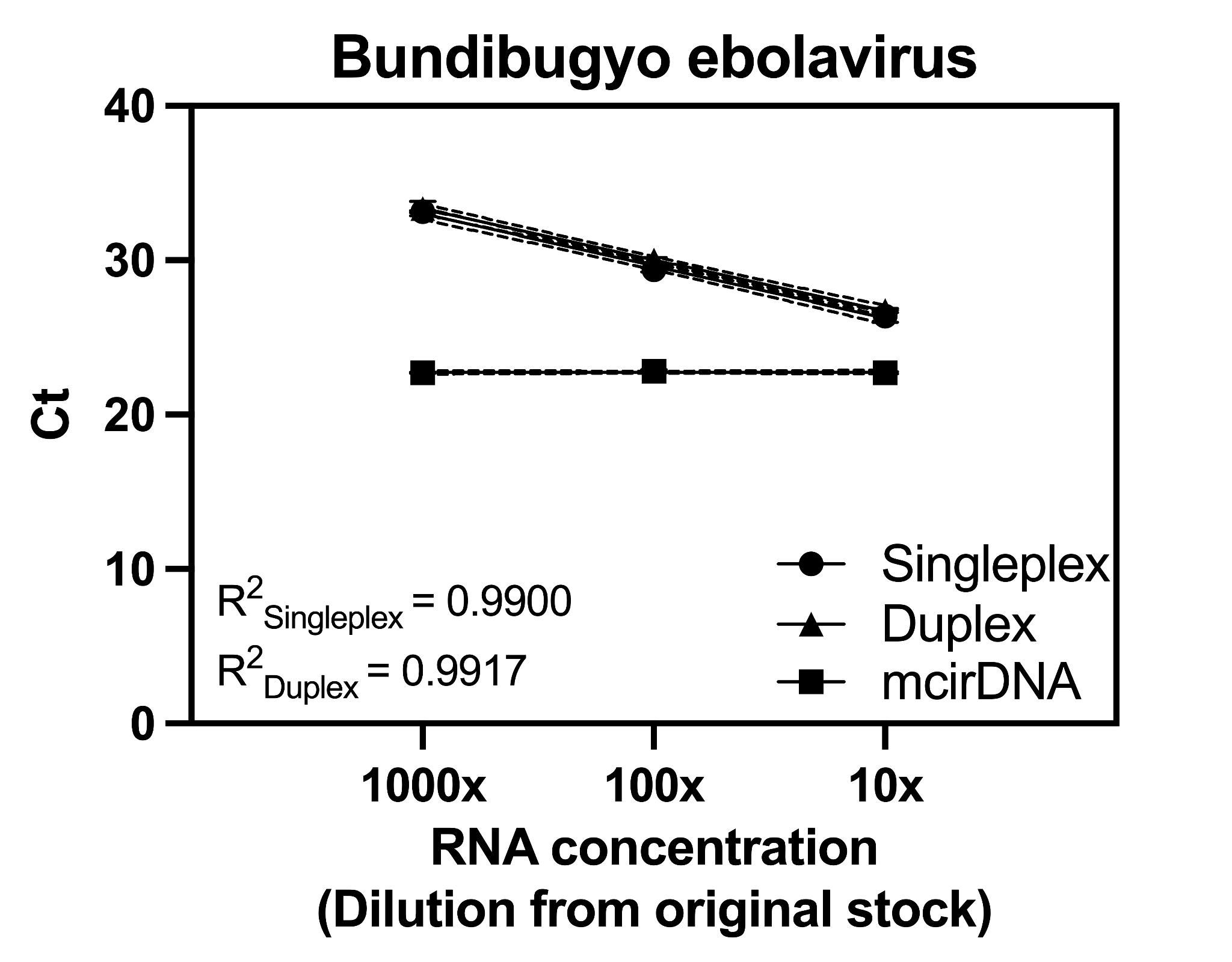

Figure A4: Detection of Bundibugyo ebolavirus (BDBV) whole viral RNA in contrived clinical samples. We generated contrived clinical samples by spiking BDBV whole viral RNA into normalized healthy human plasma background at three concentrations prepared by 10-fold serial dilutions of stock RNA. The BDBV assay showed linear detection across the dilution series in both singleplex and duplex formats. For the duplex assay, the mitochondrial circular DNA (mcirDNA) internal control showed a consistent cycle threshold (Ct) across BDBV dilutions, reflecting the constant human plasma background. Data points show the mean and standard deviation of triplicate reactions. Solid lines show simple linear regressions, and dotted lines show the 95% confidence intervals for the regressions.

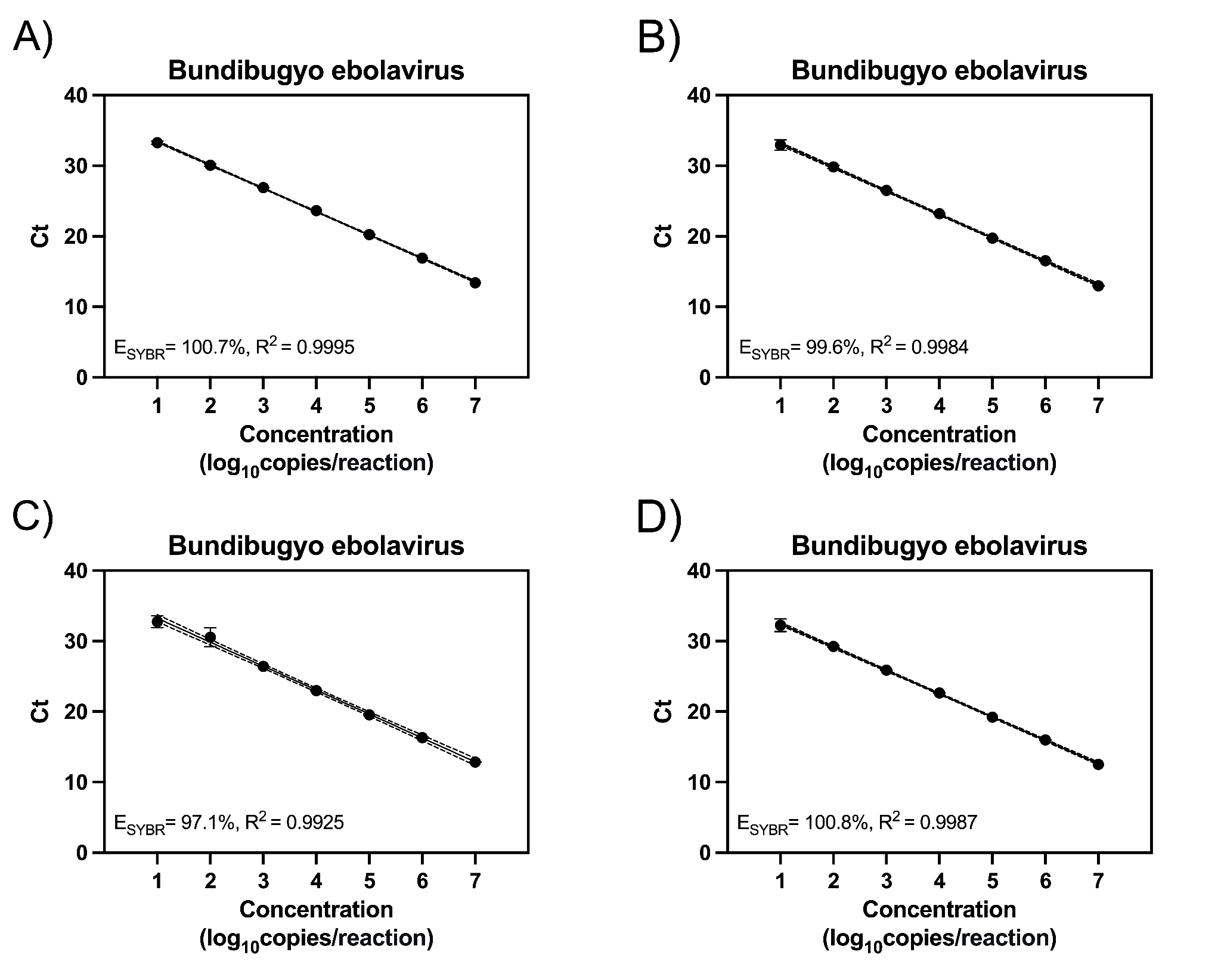

Figure A5: Optimization of primer concentration for the Bundibugyo ebolavirus qPCR SYBR Green singleplex assay. We evaluated the SYBR Green singleplex assay at the following forward and reverse primer concentrations: A) 100nM, B) 150nM, C) 200nM, D) 450nM. Standard curves were generated with synthetic RNA gene fragments. Data points represent the mean and standard deviation of triplicates. The solid line represents a simple linear regression and the dotted lines represent the 95% confidence interval of the linear regression.

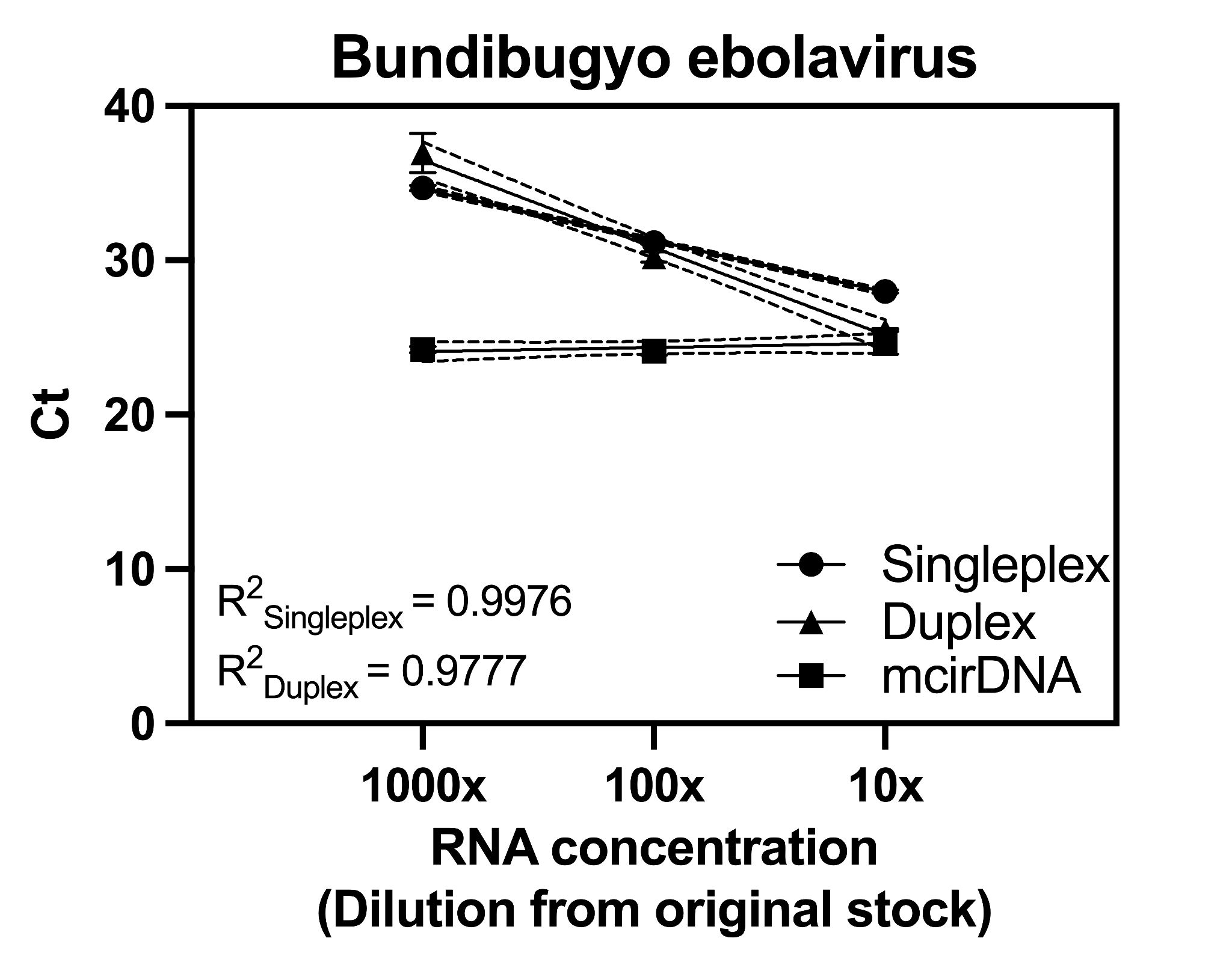

Figure A6: Detection of Bundibugyo ebolavirus (BDBV) whole viral RNA in contrived clinical samples utilizing the fast Mic qPCR cycler protocol. We generated contrived clinical samples by spiking BDBV whole viral RNA into normalized healthy human plasma background at three concentrations prepared by 10-fold serial dilutions of stock RNA. The BDBV assay showed linear detection across the dilution series in both singleplex and duplex formats. For the duplex assay, the mitochondrial circular DNA (mcirDNA) internal control showed a consistent cycle threshold (Ct) across BDBV dilutions, reflecting the constant human plasma background. Data points show the mean and standard deviation of triplicate reactions. Solid lines show simple linear regressions, and dotted lines show the 95% confidence intervals for the regressions.

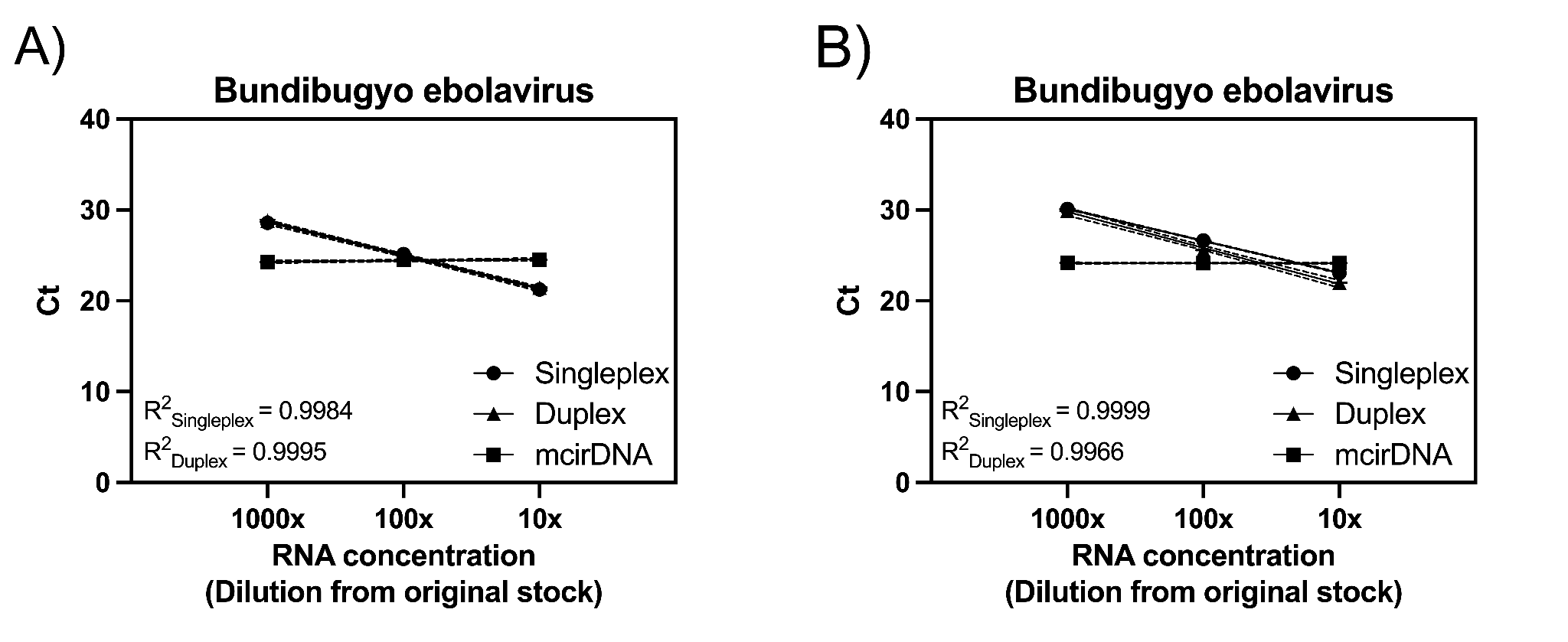

Figure A7: Detection of Bundibugyo ebolavirus (BDBV) whole viral RNA in contrived clinical samples utilizing lyophilized mastermix reagents on the QuantStudio 6 and the Mic qPCR cycler. We generated contrived clinical samples by spiking BDBV whole viral RNA into normalized healthy human plasma background at three concentrations prepared by 10-fold serial dilutions of stock RNA which were run on lyophilized RT-qPCR mastermix on both the a) QuantStudio 6 Flex and the b) Mic qPCR cycler. The BDBV assay showed linear detection across the dilution series in both singleplex and duplex formats on both instruments. For the duplex assay, the mitochondrial circular DNA (mcirDNA) internal control showed a consistent cycle threshold (Ct) across BDBV dilutions, reflecting the constant human plasma background on both instruments. Data points show the mean and standard deviation of duplicate reactions. Solid lines show simple linear regressions, and dotted lines show the 95% confidence intervals for the regressions.
